# Prospective Mendelian Randomization Study of Ancestry-Specific Blood-Cell Genetics in Predicting Pan-Cancer Risk Across 28 Malignant Neoplasms

**DOI:** 10.1101/2024.05.18.24307567

**Authors:** Jinghao Liang, Xinyi Zhou, Yijian Lin, Yuanqing Liu, Zixian Xie, Hongmiao Lin, Tongtong Wu, Xinrong Zhang, Zhaofeng Tan, Ziqiu Cheng, Weiqiang Yin, Zhihua Guo, Wenzhe Chen

## Abstract

**Background:** Research on the link between hematological characteristics and cancer risk has gained significant attention. Traditional epidemiological and cell biology studies, have identified correlations between blood traits and cancer risks. These findings are important as they suggest potential risk factors and biological mechanisms. However, these studies often can’t confirm causality, pointing to the need for further investigation to understand these relationships better.

**Methods:** Mendelian randomization (MR), utilizing single-nucleotide polymorphisms as instrumental variables, was employed to investigate hematological trait causal effects on cancer risk. Thirty-six hematological traits were analyzed, and their impact on 28 major cancer outcomes was assessed using data from the FinnGen cohort, with eight major cancer outcomes and 22 cancer subsets. Furthermore, 1,008 MR analyses were conducted, incorporating sensitivity analyses (weighted median, MR-Egger, and MR-PRESSO) to address potential pleiotropy and heterogeneity.

**Findings:** The analysis (data from 173,480 individuals primarily of European descent) revealed significant results. A decrease in eosinophil count was associated with a reduced risk of colorectal malignancies (OR 0.7702, 95% CI 0.6852, 0.8658; p = 1.22E-05). Similarly, an increase in total eosinophil and basophil count was linked to a decreased risk of colorectal malignancies (OR 0.7798, 95% CI 0.6904, 0.8808;p = 6.30E-05). Elevated hematocrit (HCT) levels were associated with a reduced risk of ovarian cancer (OR 0.5857, 95% CI 0.4443, 0.7721;p =1.47E-04). No significant heterogeneity or horizontal pleiotropy was observed.

**Interpretation:** Specific hematological traits may serve as valuable indicators and biomarkers for cancer monitoring.

**Funding:** None.

**RESEARCH IN CONTEXT:** *Evidence before this study:* Preclinical and conventional epidemiological studies have identified correlations between hematological characteristics and cancer risks. For instance, elevated eosinophil levels have been linked to improved prognosis in colorectal cancer (CRC) patients, and a high basophil-to-lymphocyte ratio (BLR) has been associated with adverse outcomes in prostate cancer. Additionally, increased red cell distribution width (RDW) has been correlated with poorer survival outcomes in metastatic penile and muscle-invasive bladder cancers. These findings suggest potential roles for hematological traits in cancer risk assessment and treatment strategies. However, traditional research methods, including randomized controlled trials (RCTs), face ethical and practical limitations, while observational studies suffer from biases and confounding variables, complicating the establishment of causal relationships.

*Added value of this study:* This study represents the first comprehensive application of Mendelian randomization (MR) to evaluate causal relationships between hematological characteristics and cancer risk. MR uses genetic variations as instrumental variables to minimize confounding, providing more reliable causal insights. Thirty-six hematological traits were analyzed, and their impact on 28 major cancer outcomes was assessed using data from the FinnGen cohort. Significant findings include the negative association between eosinophil count and CRC risk, supporting previous research on eosinophils’ antitumor role. Increased total eosinophil and basophil counts were linked to decreased CRC risk. Elevated hematocrit (HCT) levels were associated with a reduced risk of ovarian cancer, suggesting these traits could be potential targets for cancer treatment.

*Implications of all the available evidence:* Our findings provide new insights into the role of hematological traits in cancer risk, emphasizing their potential in cancer treatment and as prognostic biomarkers.

## INTRODUCTION

The global burden of cancer continues to rise, with millions of new diagnoses reported annually, highlighting the urgent need for more effective prevention, diagnosis, and treatment methods.^1^ The swift progression and high mortality rates associated with cancer underscore the importance of comprehensively understanding its pathogenesis.^2^ While significant progress has been made in immunotherapy for specific cancers such as melanoma and lung cancer, the increasing prevalence of certain cancers underscores the existing gap in preventive and treatment strategies, necessitating novel approaches.

Recent investigations have identified specific hematological characteristics as potential biomarkers for cancer detection and prognosis. For instance, elevated eosinophil levels have been linked to improved prognosis in patients with colorectal cancer (CRC).^3–5^ Research by Agreen Hadadi on patients with prostate cancer has shown that an increased basophil-to-lymphocyte ratio (BLR) is significantly associated with adverse clinical outcomes.^6^ Studies by Patel et al. and Yilmaz et al. have found that an increased red cell distribution width (RDW) correlates with poorer survival outcomes in patients with metastatic penile and muscle-invasive bladder cancers.^7,8^ Additionally, platelet count has been established as a potential prognostic marker for colorectal and endometrial cancers.^9^These findings collectively suggest a crucial role for hematological indices in assessing cancer risk and guiding therapeutic strategies.

Conventional research methodologies face significant challenges in elucidating precise mechanisms. Randomized controlled trials (RCTs), the gold standard for evaluating intervention efficacy, encounter ethical and practical limitations in cancer research.^10^ Observational studies, including case-control and cohort designs, are common but often suffer from biases and confounding variables, complicating the establishment of causal relationships.^11^ In vivo and in vitro studies, while valuable, are resource-intensive, ethically challenging, and encounter scalability issues in cancer research.

Mendelian randomization (MR) offers a distinct solution to these challenges by using genetic variations as instrumental variables to investigate causal relationships between traits and cancer outcomes. This approach minimizes confounding by leveraging the random distribution of genetic variants, providing more reliable causal insights.^12^ The validity and efficacy of MR have been demonstrated in various studies, highlighting its ability to uncover the genetic underpinnings of diseases and potential therapeutic targets.

This study utilized MR to explore the connections between 36 hematological traits and 28 major cancers, revealing significant findings such as the inverse association between eosinophil counts and colorectal cancer risk. These results not only support existing research but also enhance our understanding of the pivotal role of hematological characteristics in cancer progression. This knowledge opens up new perspectives for future cancer prevention, diagnosis, and treatment strategies.

## METHODS

### Study design

This research was conducted in accordance with the STROBE-MR guidelines, which enhance the reporting of epidemiological observations through Mendelian Randomization.^13^ A two-sample Mendelian randomization (MR) approach was used to examine the causal relationships between 36 hematological traits and 28 major cancers, utilizing genetic variants as instrumental variables (IVs). We used publicly available summary statistics from genome-wide association studies (GWAS) of hematological traits and from the FinnGen study as well as various cancer associations for cancer outcomes. All participants in the GWAS studies were of European ancestry, and there was no overlap between the groups studied for exposure and those for outcomes.

### Genetic association of hematological traits

Genetic information for these hematological traits was obtained from a comprehensive study that explored allelic variations linked to human blood traits and their connections to common complex diseases, involving 173,480 individuals of European descent from three major UK cohorts.^14^ For the cancer outcomes, data was sourced from the Finnish registry, which aided in enhancing our analysis across various cancer types and subtypes. The exposure variables involved three categories of hematological traits:White Blood Cell-related Indices:Includes counts and percentages for white blood cells, such as basophilic, eosinophilic, and other granulocytes; also includes metrics for myeloid cells, lymphocytes, and monocytes. For red blood cells, it covers total count, distribution width, hemoglobin levels, hematocrit, average volume and hemoglobin content, and reticulocyte count. Platelet-related measurements include average volume, total count, plateletcrit, and distribution width.

### Genetic associations with cancer

We specifically obtained summary statistics for 28 major cancer outcomes from the FinnGen database study.These cancer outcomes included malignancies such as tonsils and base of the tongue, bladder, bone and joint cartilage, brain, breast, bronchus and lung, cervix, colon, eye and adnexa, head and neck, intrahepatic ducts, biliary tract and gallbladder, kidney (excluding renal pelvis), nasopharynx, esophagus, oral cavity, ovaries, pancreas, prostate, rectum, small intestine, stomach, testes, thymus, thyroid, urinary organs, vulva, other skin malignancies, and primary lymphatic and hematopoietic malignancies.The FinnGen study is an ongoing cohort study that includes data from 412,181 participants (230,310 females and 181,871 males).

To enhance the robustness of our findings, we examined eight additional cancer outcomes provided by various collaborative cancer groups. We carried out further Mendelian Randomization (MR) analyses using the same exposure data and compared these results with prior MR analyses from the GWAS and FinnGen cohorts. The additional cancer outcomes included colorectal cancer data from the GWAS catalog (GCST90255675), involving 107,143 cases and 78,473 controls. These data were collected from the Cancer Interdisciplinary Studies, the Genetics and Epidemiology of Colorectal Cancer Consortium (GECCO), the Colon Cancer Family Registry (CCFR), and the UK Biobank. Renal cell carcinoma data were sourced from the genotype and phenotype database (dbGaP; Phs001736.v2.p1), including 10,784 cases and 20,406 controls from the International Agency for Research on Cancer (IARC), the National Cancer Institute (NCI), MD Anderson Cancer Center in Texas, and the Institute of Cancer Research in the UK. Prostate cancer data were from the Prostate Cancer Database and Cancer Association Group to Investigate Cancer Associated changes in Genome (PRACTICAL) consortium, comprising 79,194 cases and 61,112 controls. Ovarian cancer information from the GWAS catalog (GCST004415) included 22,406 diagnosed cases and 40,941 controls, provided by the Ovarian Cancer Association Consortium (OCAC). Cervical cancer data from the GWAS catalog (GCST012776) comprised 363 cases and 861 controls of German descent. Oropharyngeal cancer data from the GWAS catalog (GCST012241) included 1,090 cases and 2,928 controls of European descent, and oral cancer data from the GWAS catalog (GCST012237) included 1,223 cases and 2,928 controls of European descent. We also obtained data on 22 cancer subtypes, including colorectal, ovarian, stomach, vulvar, malignant melanoma, and head and neck cancers, from the Finnish database.

### Statistical analysis

Genetic variants were used as instrumental variables (IVs) in our Mendelian Randomization (MR) analysis to explore the causal effects of hematological traits on cancer and its subtypes. These variants were selected based on their strong associations with blood traits, with a significance threshold set at p<5×10^-8. For a variant to qualify as an IV, it had to meet three critical criteria: a robust association with the hematological trait, independence from confounders that affect both the blood trait and cancer outcomes, and a direct effect on the cancer outcome mediated only through the blood trait.^12^

To manage linkage disequilibrium (LD) among single nucleotide polymorphisms (SNPs), which is the non-random association of alleles, it was essential to ensure independence among SNPs before conducting MR analysis. LD was addressed using the TwoSample MR software package, setting parameters to r ² = 0.001 and kb to maintain SNP independence.^15^ The strength of each SNP’s association was determined using the F-statistic, calculated by R²= (2β²× EAF × (1 -EAF))/(2β²× EAF × (1 -EAF)+ 2N × EAF × (1 -EAF) × SE²) and F = (R²× (N -2))/(1 -R²), where EAF is the effect allele frequency, β and SE are the effect size and standard error, respectively, and N is the sample size for the hematological trait.^14^ SNPs with an F-statistic below 10 were excluded as weak instrumental variables.^16^

We executed Mendelian randomization (MR) analysis across all exposure-outcome combinations, encompassing 1008 tests (36 exposure variables multiplied by 28 cancer outcomes from the FinnGen data).To synthesize these relationships, we employed the random-effects inverse-variance weighted (IVW) method, which assumes the validity of all genetic instruments. However, this method can be biased when a significant proportion of the instrumental variables (IVs) exhibit pleiotropy.^17^ To address the inherent issue of multiple testing in our analysis and to reduce the risk of Type I errors, we applied Benjamini-Hochberg false discovery rate (FDR) correction during the ’Validation’ and ’Discovery’ phases. We set the threshold for FDR-adjusted P-values (q-values) at <0.05 to define “strong evidence” of causality. Additionally, results with q-values between ≥0.05 and <0.20 were classified as providing “suggestive evidence,” indicating that potential associations might require further investigation.^18^

In addressing potential pleiotropy among IVs, we utilized MR-PRESSO global test, outlier test, and distortion test to identify and exclude SNPs exhibiting such effects. When outliers were detected, we re-evaluated the causal relationships to maintain the integrity of our findings. MR-Egger intercept tests, along with Cochran’s Q test and MR-Egger model within the IVW framework, played a crucial role in assessing pleiotropy and heterogeneity in our analysis. In the presence of pleiotropy, we particularly favored the MR-Egger method, while in cases where Cochran’s Q test indicated significant heterogeneity (P<0.05), we opted for the weighted median (WM) model for a more detailed analysis.^19,20^ Conversely, in the absence of significant heterogeneity, we employed a fixed-effect model. Additionally, we conducted leave-one-out sensitivity analysis to verify the robustness of our results.

## RESULTS

Our MR study meticulously evaluated 36 hematological traits, each supported by a substantial array of instrumental variables (IVs) ranging from 59–243. F-statistics for these IVs varied from 29.7–2806.4, exceeding the commonly recommended threshold of 10, indicating strong instrumental validity and mitigating concerns about weak instrument bias (details in Supplementary files).

### Analysis of FinnGen cancer outcomes

Meta-analyses included 1,008 associations across 36 hematological traits and 28 cancer outcomes. The IVW analysis identified 83 statistically significant associations (Fig. 1). After adjusting for multiple testing with the Benjamini-Hochberg false discovery rate (FDR) correction, three key associations emerged with strong evidence: decreased risk of CRC associated with increased eosinophil counts (OR 0.7702, 95% CI 0.6852,0.8658; p = 1.22E-05), reduced risk of CRC associated with combined eosinophil and basophil counts (OR 0.7798, 95% CI 0.6904,0.8808; p = 6.30E-05) and lower risk of ovarian cancer associated with higher HCT levels (OR 0.5857, 95% CI 0.4443, 0.7721; p = 1.47E-04). Details are provided in Figure 3 and Supplementary File 1: Tables S1–S3.These results were corroborated by a robust set of instrumental variables (IVs) with 159, 157, and 108 IVs showing low variability (I²=18.24%, I²=20.35%, and I²=3.59%, respectively). No signs of pleiotropy were observed using the MR-Egger intercept or MR-PRESSO diagnostic tools, and no anomalous SNPs were detected. The findings maintained their statistical significance in the weighted median analysis, which suggests stability despite potential horizontal pleiotropy in some IVs. Examination of funnel and scatter plots did not indicate any noticeable bias or inconsistency. Among the 1,008 associations tested, 494 had OR values less than 1, whereas the others had OR values greater than 1, as shown in Fig. 2.

**Fig. 1:**
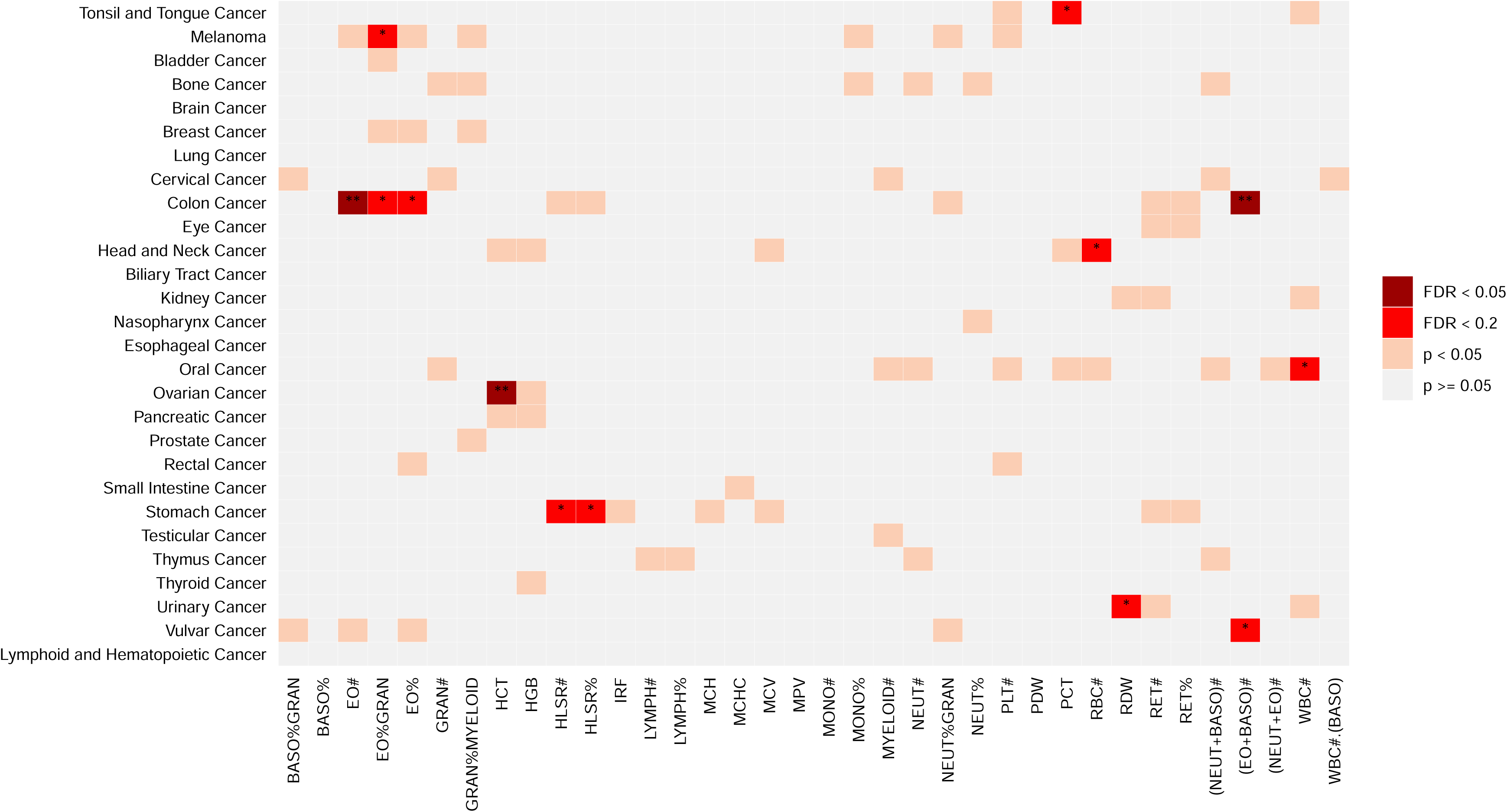
Results of Mendelian randomization associations of hematological traits with cancer outcomes from FinnGen cohort. * A false discovery rate (FDR)-corrected P-value (“q-value”) 0.05 ≤ q-value < 0.20 to define “suggestive evidence” ** A false discovery rate (FDR)-corrected P-value (“q-value”) <0.05 was used as a threshold to define “strong evidence”

**Fig. 2:**
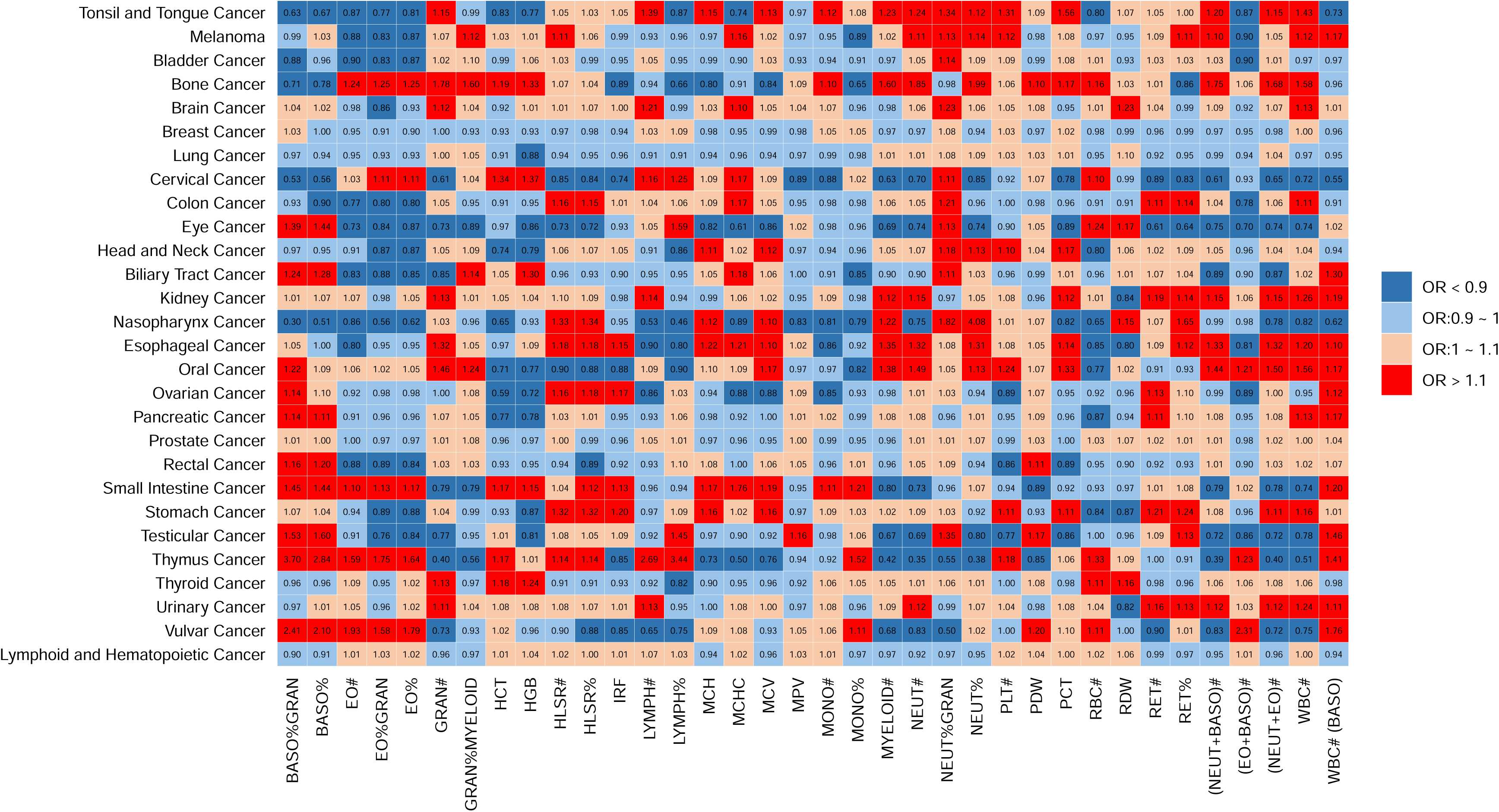
In the context of Mendelian randomization analysis, the relationships between 36 blood cell types and cancer risk were quantified using odds ratios (ORs). An OR greater than 1 indicates that alterations in specific blood cell types are potentially associated with an increased risk of cancer. Conversely, an OR less than 1 suggests that changes in these hematological traits may confer a protective effect, potentially reducing the risk of cancer. This statistical approach provides insight into how variations in blood cell populations can influence cancer susceptibility.

**Fig. 3:**
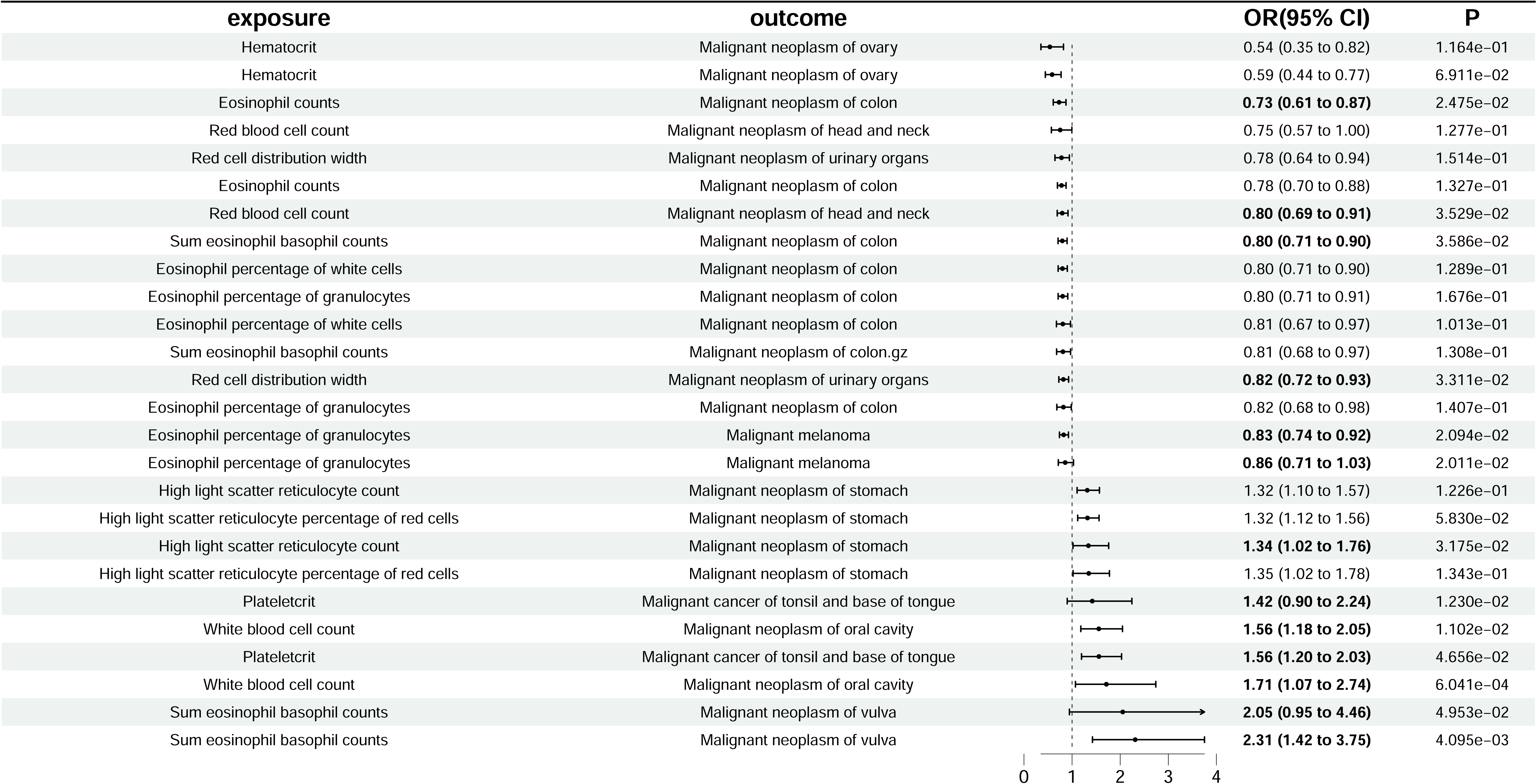
Statistically significant associations between hematological traits and cancer outcomes in FinnGen meta-analysis. Associations are listed in ascending order of p-values in the inverse variance-weighted model. The forest plot represents the pooled odds ratio under the inverse variance-weighted model.Association with a false discovery rate (FDR) -corrected P-value (“q-value”) <0.20 are shown here. Abbreviations: CI, confidence interval; NSNP, number of single-nucleotide polymorphisms.

Following robust relationships supported by strong evidence, we now focus on those supported by suggestive evidence. An inverse relationship was observed between colon cancer and both eosinophil percentage in white blood cells (OR 0.801, 95% CI 0.711, 0.901; p =2.31E-04) and total eosinophil and basophil counts (OR 0.804, 95% CI 0.7101, 0.911; p = 6.08E-04). Likewise, a higher proportion of eosinophils in granulocytes correlated with a lower likelihood of developing malignant melanoma (OR 0.8258, 95% CI 0.7375-0.9247; p = 0.000908). Conversely, increased counts of eosinophils and basophils were positively linked to the risk of vulvar malignancies (OR 2.312, 95% CI 1.425-3.753; p = 0.000693). A rise in plateletcrit was associated with a heightened risk of cancers affecting the tonsils and the base of the tongue (OR 1.558, 95% CI 1.196-2.029; p = 0.001). Additionally, a higher percentage of high light scatter reticulocytes was connected to an elevated risk of stomach cancer (OR 1.32, 95% CI 1.117-1.567; p = 0.001), while high light scatter reticulocyte counts were positively correlated with the risk of malignant stomach neoplasms (OR 1.316, 95% CI 1.104,1.569; p = 0.002). Conversely, an increased red blood cell count correlated with a decreased risk of head and neck cancer (OR 0.7957, 95% CI 0.6921,0.9149; p = 0.001), and red cell distribution width was negatively associated with the risk of malignant neoplasms of the urinary organs (OR 0.82, 95% CI 0.725,0.928; p = 0.002). Additionally, an increased white blood cell count was associated with an increased risk of malignant neoplasms of the oral cavity (OR 1.556, 95% CI 1.181,2.048; p = 0.002), as shown in Fig. 3 and Supplementary Files S4–S13.

In our MR analysis, horizontal pleiotropy was detected in the relationship between red blood cell count and oral cavity cancer (MR-PRESSO p=0.021). However, the association remained statistically significant even after removing outlier SNPs (OR=0.77, 95% CI: 0.61–0.97, p=0.0278, MR-PRESSO p=0.066). Similarly, horizontal pleiotropy was observed in the association between platelet count and breast cancer (MR-PRESSO p=0.024). However, this association persisted after excluding outliers (OR=1.028, 95% CI: 0.968–1.093, p=0.0278; MR-PRESSO p=0.134).

### Analyses of cancer subtypes

The analysis of 28 exposures across 22 cancer subtypes identified 176 statistically significant associations. Notably, consistent associations were observed across major cancer types and subtypes, including three colorectal malignancies (colorectal adenocarcinoma, CRC, and colorectal adenocarcinoma), ovarian cancers (ovarian serous carcinoma and ovarian endometrioid carcinoma), gastric cancer subtypes (adenocarcinoma and papillary adenocarcinoma of the stomach), and subtypes of malignant melanoma. Four hematological markers associated with eosinophil count (eosinophil count, total eosinophil and basophil counts, percentage of eosinophils among leukocytes, and percentage of eosinophils among granulocytes) were inversely related to the risk of colorectal malignancies. Notably, the risk of colorectal adenocarcinoma was significantly inversely correlated with eosinophil count (OR 0.756, 95% CI 0.663,0.862; p = 3.07E-05), total eosinophil count, basophil count (OR 0.774, 95% CI 0.676,0.885; p = 1.86E-04), eosinophil percentage of leukocytes (OR 0.788, 95% CI 0.691,0.899; p = 4.25E-04), and eosinophil percentage of granulocytes (OR 0.796, 95% CI 0.691,0.917; p = 0.002). This negative association was consistent across different CRC subtypes. Similarly, ovarian serous carcinoma (OR 0.652, 95% CI 0.484,0.878; p = 0.005) and ovarian endometrioid carcinoma (OR 0.521, 95% CI 0.280,0.969; p = 0.039) showed consistent negative associations with HCT levels.

Additionally, adenocarcinoma and papillary adenocarcinoma of the stomach were positively correlated with a high light scatter reticulocyte percentage of red blood cells (OR 1.44, 95% CI 1.155,1.795; p = 0.001) and malignant melanoma of the skin was negatively associated with an eosinophil percentage of granulocytes (OR 0.832, 95% CI 0.728,0.951; p = 0.007), consistent with their primary tumor profiles.

### Analyses of consortia cancer outcomes

Of the 288 cancer-related associations analyzed (36 exposures × 8 cancer outcomes), 95 were statistically significant(Fig. 4). Among these, five associations showed a significant inverse correlation between CRC risk and eosinophil-related markers, including eosinophil count (OR 0.893, 95% CI 0.834,0.957; p = 0.001), eosinophil percentage of granulocytes (OR 0.896, 95% CI 0.831,0.965; p = 0.004), eosinophil percentage of leukocytes (OR 0.886, 95% CI 0.825,0.952; p = 9.31E-04), and total count of eosinophils and basophils (OR 0.904, 95% CI 0.846,0.965; p = 0.003).

**Fig. 4:**
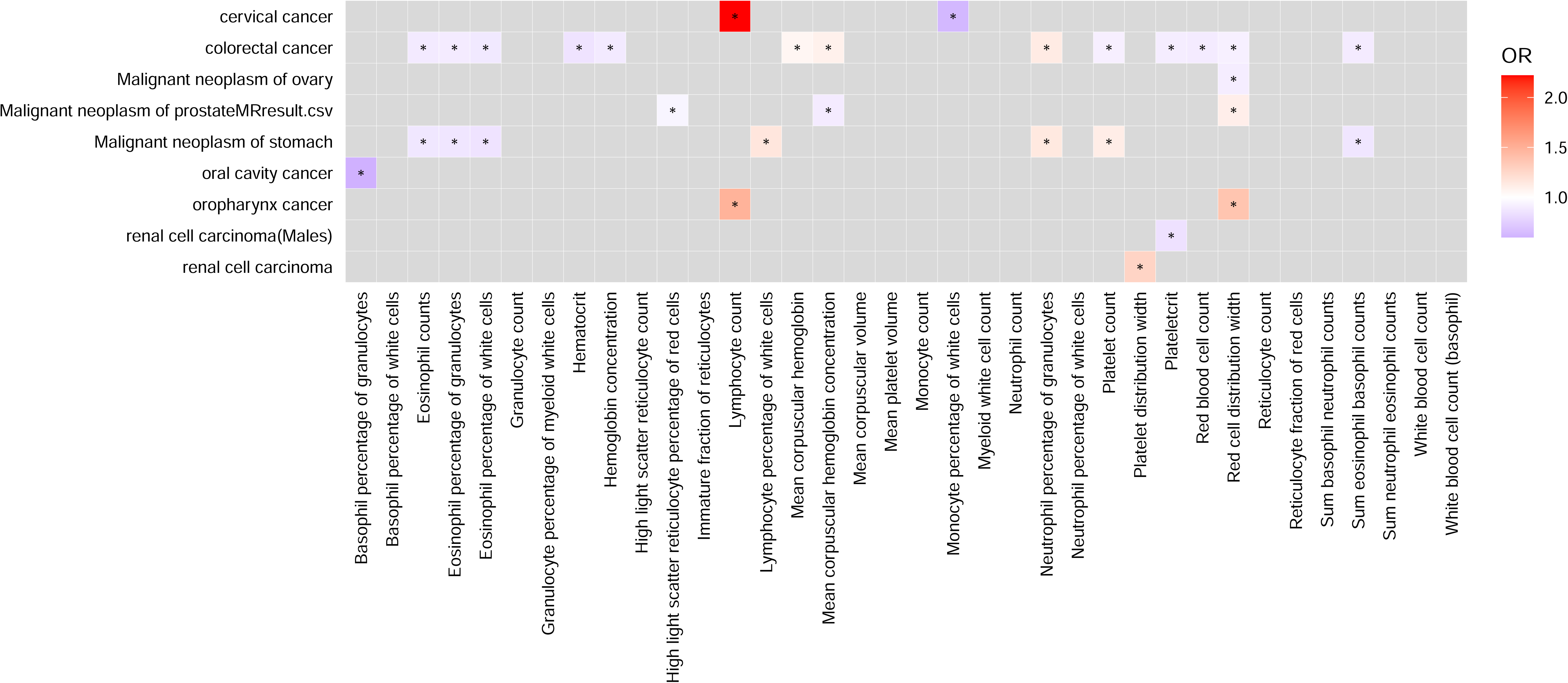
Results of Mendelian randomization associations of hematological traits with cancer outcomes from combined GWAS and various cancer consortiums. The relationships between 36 hematological traits and cancer risk were quantified using odds ratios (ORs). An OR greater than 1 indicates a potential increased cancer risk, while an OR less than 1 suggests a protective effect, reducing cancer risk * P < 0.05, indicating that the associations are statistically significant at the 95% confidence level. This means that changes in these specific hematological traits are likely associated with cancer risk.

Additionally, an increased risk of prostate cancer was positively associated with RDW (OR 1.1, 95% CI 1.05,1.15; P = 9.03E-05). These findings had similar effect sizes and statistical significance compared with the analyses conducted on cancer associations and in the FinnGen study.

## DISCUSSION

This study represents the first comprehensive application of MR to evaluate causal relationships between hematological characteristics and the risk of various cancers. Using extensive genomic association study data on blood cells and reliable cancer data from Finland, we analyzed 36 hematological traits and their relationships with 28 different cancers. This systematic approach confirmed the significant role of these three specific hematological traits in the etiology of the two cancers.

One of the most robust findings was the significant negative association between eosinophil count and CRC risk. This observation aligns with previous research demonstrating a close association between eosinophilic infiltration and a more favorable prognosis in patients with CRC.^3–5^ Eosinophils can release various bioactive substances, such as eosinophil cationic protein (ECP), tumor necrosis factor alpha (TNFα), and proteolytic granzyme A, potentially inhibiting CRC growth and spread by directly killing tumor cells or modulating immune responses within the tumor microenvironment.^21^Additionally, by altering the Th1/Th2 immune response balance and promoting the activation and aggregation of effector immune cells, such as T cells, eosinophils may enhance immune surveillance and attack CRC.^5^ Tumor cells can even recruit eosinophils by secreting attractants, such as eotaxin and interleukin (IL)-33, highlighting their potential role in anti-tumor immunity through degranulation.^22^ Furthermore, tumor-associated eosinophils in CRC are usually linked to a lower tumor grade and fewer metastases,^23^ particularly in patients with lymph node metastasis, showing an increased presence of lymph node eosinophils in approximately 27% of cases.^24^ These observations suggest a significant antitumor role for eosinophils in the tumor microenvironment, potentially independent of CD8+ T cells.^23^ These findings provide a strong theoretical basis for future immune therapeutic strategies targeting eosinophils and emphasize the importance of exploring and utilizing eosinophil mechanisms in CRC to develop new antitumor treatments. In our analysis of the total counts of eosinophils and basophils in the blood relative to CRC, we observed a negative association between these two factors. Regarding basophils, studies indicate that their high levels are associated with better overall survival and disease-free survival in patients with CRC,^25^ suggesting they may suppress tumor progression by secreting histamine^26^and pro-inflammatory cytokines, such as TNFα, IL-6, and IL-1β, thereby enhancing the recruitment of specific CD8+ T cells and promoting cancer cell apoptosis.^27^ Our findings imply that basophils may play a key role in regulating immune responses and suppressing CRC and that their levels are negatively correlated with the prognosis of patients with CRC. In patients with ovarian cancer, the negative association between low HCT and poor prognosis may involve multiple factors. A low HCT level often indicates anemia,^28,29^ possibly because of cancer’s high iron demand, leading to iron deficiency.^30^ Anemia can cause tissue hypoxia, which can increase tumor cell resistance to chemotherapy drugs and potentially promote tumor aggressiveness.^31,32^ Consequently, low HCT levels may reflect the severity of ovarian cancer and its response to treatment, thus affecting patient survival rates.^33^This relationship indicates that HCT levels could be a significant biomarker for evaluating the prognosis of patients with ovarian cancer.

Following the discussion of strongly supported findings, we present suggestive evidence. Similar to the strong association with CRC, we observed a significant negative correlation between the proportion of eosinophils in white blood cells and granulocytes and the risk of CRC, further emphasizing the antitumor properties of eosinophils in this context. Their antitumor activity was also reflected in the negative association observed between malignant melanoma and the percentage of eosinophils. Eosinophils can release various cytotoxins, such as major basic protein (MBP) and TNF-α, which can directly cause tumor cell dissolution.^21^Additionally, eosinophils attract CD8+ T cells by releasing chemokines, such as CCL5, CXCL9, and CXCL10, enhancing the tumor-killing activity of these cells.^34,35^ These mechanisms indicate that eosinophils play an important role in immune surveillance within the tumor microenvironment, potentially reducing the occurrence of tumors such as malignant melanoma.

However, eosinophils also exhibit pro-tumor activity in certain cancers. For example, we found a positive association between malignant neoplasms of the vulva and total eosinophil and basophil counts. This link implies that eosinophils and basophils could facilitate tumor development by influencing the tumor microenvironment and enhancing the migration and proliferation of cancer cells. Notably, eosinophils can promote local accumulation of tumor cells by secreting chemokines, such as CCL6,^36^ without altering the survival capability of these cells. Additionally, basophils have been identified as potential biomarkers in tumor research. For instance, Agreen Hadadi’s research on a group of patients with metastatic hormone-sensitive prostate cancer (mHSPC) demonstrated that a higher basophil-to-lymphocyte ratio (BLR) was significantly linked to worse clinical outcomes.^6^ Studies on the role of basophils in bladder and pancreatic cancers suggest that an increase in these cells is associated with tumor recurrence and poor survival prognosis,^37,38^ possibly owing to the release of pro-inflammatory and growth-promoting factors in the tumor microenvironment. These findings highlight the importance of understanding the specific functions and mechanisms of eosinophils and basophils in malignant vulvar tumors and the necessity of considering them as potential therapeutic targets or disease monitoring indicators.

Our study also identified associations between other hematological markers and various cancers. The positive association observed between white blood cell count and oral malignant tumors might be because of the predominance of neutrophils in the microenvironment of these tumors. Neutrophils can release cytokines, chemokines, and growth factors that promote angiogenesis, tumor cell proliferation, and migration, thus accelerating tumor growth and spread.^39^ This association suggests a potential role for white blood cell count as a biomarker for the clinical progression of oral malignant tumors, as corroborated by studies demonstrating adverse clinical outcomes associated with an increased total white cell count.^40–42^ This finding emphasizes the potential importance of white blood cell count as a biomarker for the clinical progression of oral malignant tumors. Furthermore, eosinophils and basophils exhibit dual roles in cancer development; they show antitumor properties in some cancers, while they may promote tumor development in other types, such as malignant vulvar tumors. Additionally, an increased total white blood cell count is associated with tumor progression, highlighting its importance as a potential biomarker.

Anemia is a common complication in patients with cancer, affecting overall health and treatment outcomes. For instance, studies have indicated that low levels of deglycating enzymes in red blood cells may be involved in the malignant transformation of the colonic mucosa.^43^Additionally, a retrospective study involving 758 patients demonstrated that a reduction in red blood cell count post liver cancer surgery is indicative of poorer survival rates.^44^ Similarly, in head and neck malignancies, a lower red blood cell count may suggest a poor health status, potentially reflecting more severe disease characteristics or higher metabolic demands of the tumor. This also explains the observed negative correlation between red blood cell count and malignant neoplasms of the head and neck. A high percentage of highly scattered reticulocytes linked to malignant gastric tumors indicates increased red cell production, which is typically a physiological response to chronic bleeding or iron deficiency. Therefore, chronic bleeding from gastric tumors may lead to iron deficiency anemia, stimulating the bone marrow to increase reticulocyte production to compensate for red blood cell loss, reflecting disease progression and tumor-related pathological processes. In contrast, we found a negative association between urinary system malignancies and RDW. This association may be owing to low RDW, typically indicating a higher uniformity of red blood cells, suggesting lower levels of inflammation and oxidative stress,^45^ which play a promoting role in tumor development. For instance, Patel et al. found in their study that a higher RDW was associated with a poorer overall survival rate in patients with metastatic penile cancer undergoing chemotherapy.^7^ Similarly, Yilmaz et al. showed that a higher RDW is associated with a poorer prognosis in muscle-invasive bladder cancer.^8^ These studies suggest that RDW might serve as an indirect indicator of inflammation and tumor biological activity, with a low RDW possibly associated with a better prognosis in urinary system tumors. These findings emphasize the importance of RDW as a potential prognostic indicator in patients with urinary tumors. Changes in red cell parameters are related to the presence and progression of tumors. Notably, high-scatter reticulocytes indicate a potential pathological state, whereas a low RDW is associated with a better prognosis in urinary system tumors, demonstrating its potential as a prognostic tool.

The positive relationship between malignant cancers of the tonsils and base of the tongue and plateletcrit may be explained by the multiple mechanisms platelets play in tumor development. Platelets can provide mechanical protection to tumor cells during circulation, enrich their bioactivity in the tumor microenvironment, and promote the transport and release of tumor molecules.^46^ Additionally, cell factors and growth factors present in platelets, such as vascular endothelial growth factor (VEGF) and epidermal growth factor (EGF),^47–50^ act as signals promoting tonsil and base of tongue cancer, facilitating invasion and metastasis. Platelet activity is crucial in facilitating tumor invasion and metastasis, especially by enhancing angiogenesis and shielding tumor cells from harm. This emphasizes the importance of platelets as potential therapeutic targets in these cancers.

A significant strength of this study is that, compared with traditional observational studies, the MR method, which uses genetic variations as instrumental variables, effectively minimizes potential biases from confounding factors and reverse causality, thereby providing clearer and more reliable evidence of causal relationships.

Furthermore, the extensive scope of this research, analyzing over 2,000 potential causal associations across multiple cancer datasets from the UK Biobank, FinnGen, and various cancer consortia, strengthens the generalizability of the findings.

Additionally, the use of multiple MR methods, including IVW, weighted median approach, MR-Egger regression, MR-PRESSO, and weighted mode approach, enhances the robustness of the research and effectively handles and confirms the directionality of causal relationships, even in cases of potential assumption violations. Moreover, this study used a large number of SNPs, improving the strength and coverage of genetic tools and enhancing the precision and reliability of causal inference.^51^ By integrating hematological characteristics with genetic data, this study not only provides a deep understanding of the relationships between hematological characteristics and cancer but also emphasizes the long-term association between lifelong exposure to hematological traits and cancer risk, an association that might not be affected by short-term interventions,^52^thus providing a solid scientific foundation for understanding the role of various blood components in cancer development.

This study has some limitations. Our analysis assumed no gene-environment or gene-gene interactions, potentially overlooking the significant roles of these factors in cancer development, particularly since the responses of eosinophils and basophils may be significantly affected by environmental factors, such as infection and inflammation. Additionally, the assumption of linear and constant effects of biomarkers may be overly simplistic and may not fully reflect the diverse roles of eosinophils across different cancer stages and types.^53^ Furthermore, MR is primarily designed to assess the long-term effects of genes on disease risk and may not be entirely applicable for evaluating the impact of short-term medical interventions on these traits.^54^ Our findings are based on European populations, and their generalizability to non-European groups remains uncertain. Lastly, limited sample sizes for rare cancers and specific cancer subtypes may have limited the statistical power to detect subtle genetic effects. While sensitivity analyses were conducted, they relied on assumptions that could not be directly verified, introducing some uncertainty in interpreting the results.

In conclusion, our study demonstrated the different possible roles and complex mechanisms underlying hematological characteristics in various cancers, such as the antitumor properties of eosinophils in CRC and their potential to promote tumor development in malignant vulvar tumors. This highlights the context-dependent effects of these cells, likely influenced by specific cytokines and chemical signals within the tumor microenvironment. These findings underscore the need to consider additional microenvironmental factors when interpreting relationships between hematological characteristics and cancer, adding complexity to future research endeavors.

## CONTRIBUTIONS

Jinghao Liang, Zhihua Guo, Wenzhe Chen designed the research, contributed to the discussion, reviewed and edited the manuscript, and takes full responsibility for the work as a whole. Xinyi Zhou, Yijian Lin, Yuanqing Liu designed the study, performed analysis, wrote the manuscript and contributed to the discussion. Zixian Xie, Hongmiao Lin, Tongtong Wu, and Xinrong Zhang conducted the research, analyzed the data, and reviewed and edited the manuscript. Zhaofeng Tan, Ziqiu cheng, Weiqiang Yin conducted the research and contributed to the discussion.

## Data Availability

All data produced in the present study are available upon reasonable request to the authors
All data produced in the present work are contained in the manuscript
All data produced are available online at

## ACKNOWLEDGMENTS

We gratefully acknowledge the participation of all UK Biobank, NIHRCambridge and BioResource.We want to acknowledge the participants and investigators of the FinnGen study Colorectal cancer: We are particularly grateful to all parties that provided valuable data resources for colorectal cancer research. The Colorectal Cancer Genetics and Epidemiology Consortium (GECCO), the Colon Cancer Family Registry (CCFR), and the UK Biobank have laid a solid foundation for our study.

Renal cell cancer: For the renal cell carcinoma research component, we would like to express our gratitude to the institutions involved in data collection and organization, namely the International Agency for Research on Cancer (IARC), the National Cancer Institute (NCI), and The University of Texas MD Anderson Cancer Center, as well as the Institute of Cancer Research, UK.

Ovarian cancer: We express our gratitude to the Ovarian Cancer Association Consortium for providing essential data support that significantly contributed to our research.

Prostate cancer: We extend our heartfelt gratitude to the institutions that supported the genome-wide association analyses for prostate cancer. Our special thanks go to the Canadian Institutes of Health Research, the European Commission’s Seventh Framework Programme (Grant Agreement No. 223175, Project Name HEALTH-F2-2009-223175), Cancer Research UK (Grants C5047/A7357, C1287/A10118, C1287/A16563, C5047/A3354, C5047/A10692, and C16913/A6135), and the National Institutes of Health (NIH) Cancer Post-Cancer GWAS initiative (Grant No. 1 U19 CA 148537-01, the GAME-ON initiative).

Gastric cancer: We used gastric cancer data provided by the BioBank Japan Project in our research, and we are grateful for the contributions of the project and all participants.

Oral cavity and Oropharynx: We are grateful to the International Head and Neck Cancer Epidemiology Consortium (INHANCE), the European Prospective Investigation into Cancer and Nutrition (EPIC), and the HN5000 project teams for providing valuable data support that facilitated our research.

## DATA SHARING STATEMENT

Data sources are detailed in Supplementary Table S1. Data are available upon reasonable request. A limited dataset may be available upon reasonable request.

