## Supplementary for "Prospective Mendelian Randomization Study of Ancestry-Specific Blood-Cell Genetics in Predicting Pan-Cancer Risk Across 28 Malignant Neoplasms"

Authorship

Jinghao Liang^1^, Xinyi Zhou^2^ , Yijian Lin^2^, Yuanqing Liu^2^, Zixian Xie^1^, Hongmiao Lin^3^, Tongtong Wu^2^, Xinrong Zhang^2^, Zhaofeng Tan^3^, Ziqiu Cheng^1^, Weiqiang Yin^1^, Zhihua Guo^1,*^, Wenzhe Chen^3,*^

^1^Department of Thoracic Surgery, The First Affiliated Hospital of Guangzhou Medical University, Guangzhou 510120, China

^2^Second Clinical Medical College, Guangdong Medical University, Dongguan 523000, China

^3^Graduate School, The Sixth Affiliated to Guangzhou Medical University, The Sixth People's Hospital, Guangzhou 510120, China

*Corresponding author: Zhihua Guo, Department of Thoracic Surgery, The First Affiliated Hospital of Guangzhou Medical University, Guangzhou 510120, China, E-mail address:; Wenzhe Chen, Graduate School, The Sixth Affiliated to Guangzhou Medical University, The Sixth People's Hospital, Guangzhou 510120, China, E-mail address:.

ABSTRACT

Background:

Research on the link between hematological characteristics and cancer risk has gained significant attention. Traditional epidemiological and cell biology studies, have identified correlations between blood traits and cancer risks. These findings are important as they suggest potential risk factors and biological mechanisms. However, these studies often can't confirm causality, pointing to the need for further investigation to understand these relationships better.

Methods:

Mendelian randomization (MR), utilizing single-nucleotide polymorphisms as instrumental variables, was employed to investigate hematological trait causal effects on cancer risk. Thirty-six hematological traits were analyzed, and their impact on 28 major cancer outcomes was assessed using data from the FinnGen cohort, with eight major cancer outcomes and 22 cancer subsets. Furthermore, 1,008 MR analyses were conducted, incorporating sensitivity analyses (weighted median, MR-Egger, and MR-PRESSO) to address potential pleiotropy and heterogeneity.

Findings:

The analysis (data from 173,480 individuals primarily of European descent) revealed significant results. A decrease in eosinophil count was associated with a reduced risk of colorectal malignancies (OR 0.7702, 95% CI 0.6852, 0.8658; p = 1.22E-05). Similarly, an increase in total eosinophil and basophil count was linked to a decreased risk of colorectal malignancies (OR 0.7798, 95% CI 0.6904, 0.8808;p = 6.30E-05). Elevated hematocrit (HCT) levels were associated with a reduced risk of ovarian cancer (OR 0.5857, 95% CI 0.4443, 0.7721;p =1.47E-04). No significant heterogeneity or horizontal pleiotropy was observed.

Interpretation:

Specific hematological traits may serve as valuable indicators and biomarkers for cancer monitoring.

Funding:

None.

KEYWORDS: Hematological traits, Cancer risk, Mendelian randomization, Biomarkers

Supplementary Figure 1. Genetic association of Eosinophil counts with Malignant neoplasm of colon

1) Forest plot

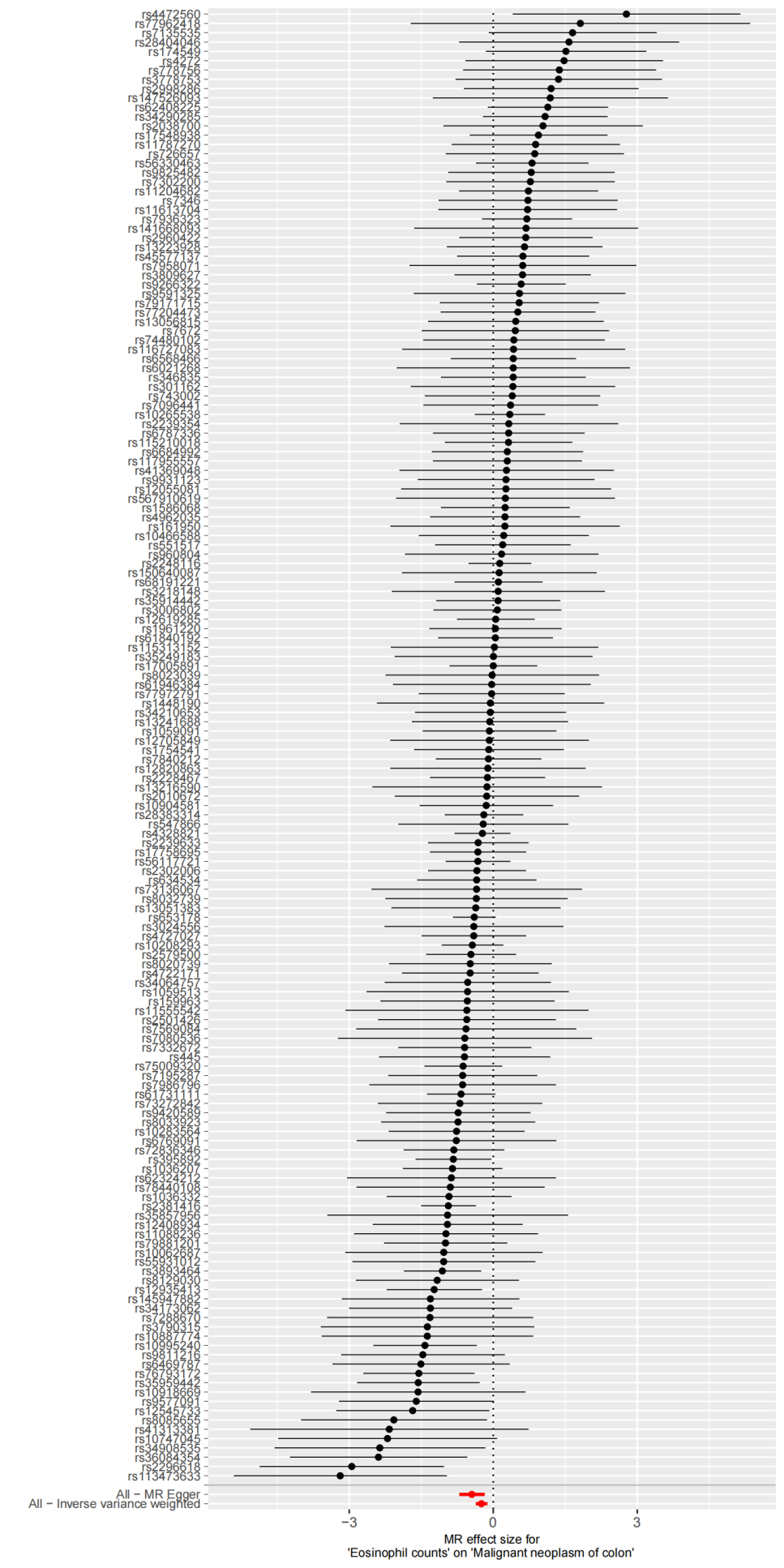

2) Leave-one-out plot

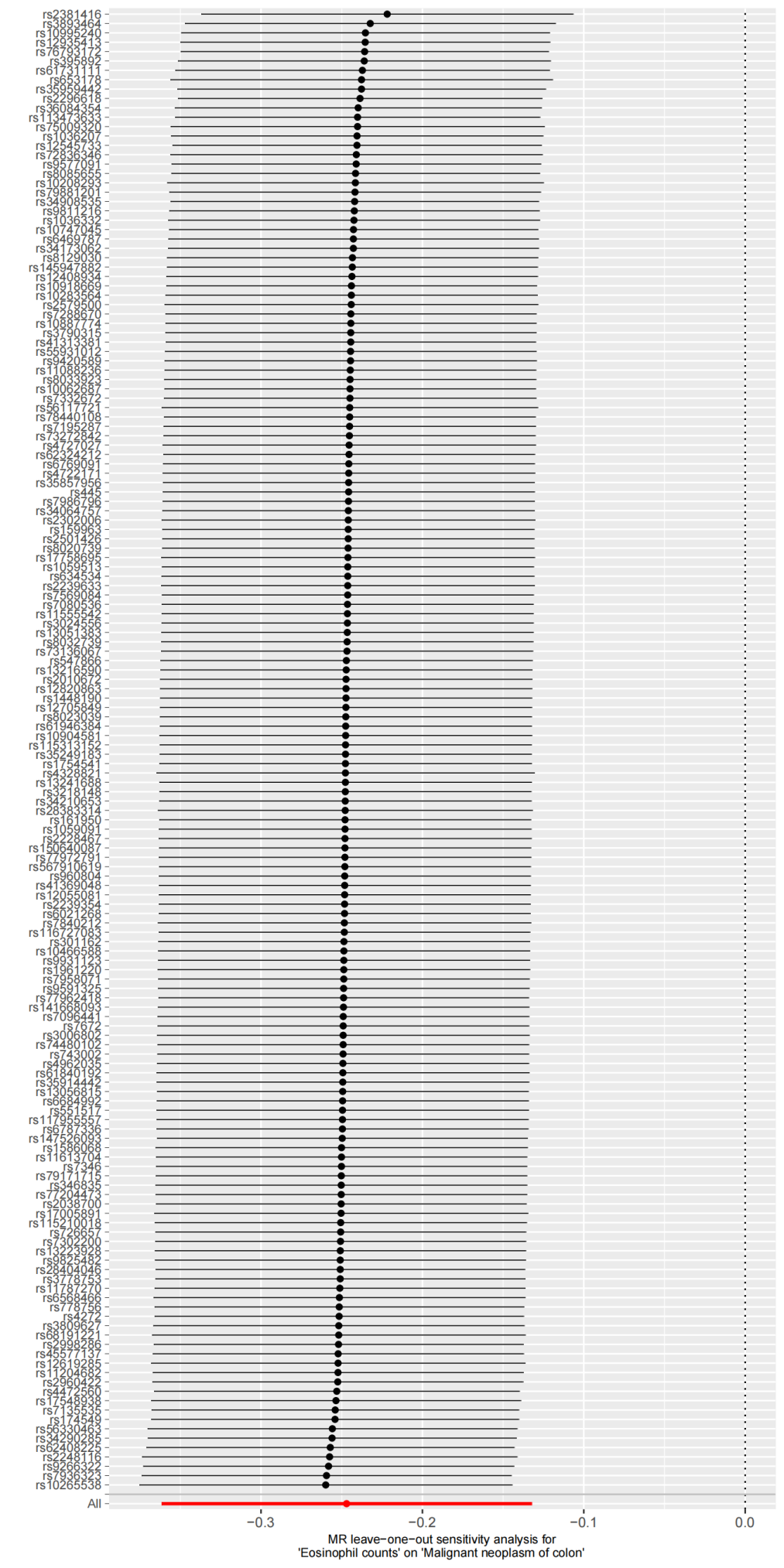

3) Scatter plot

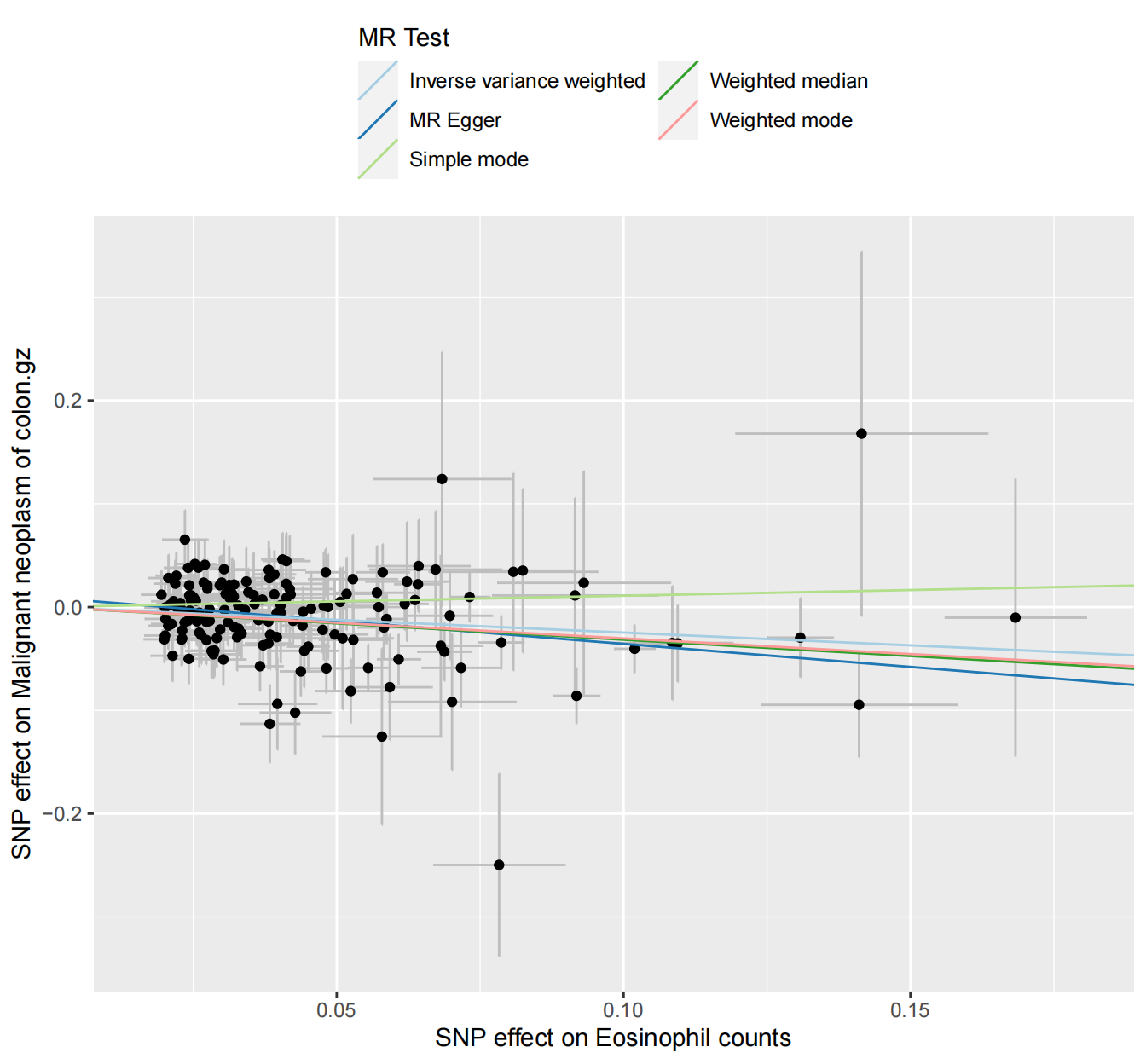

4) Funnel plot

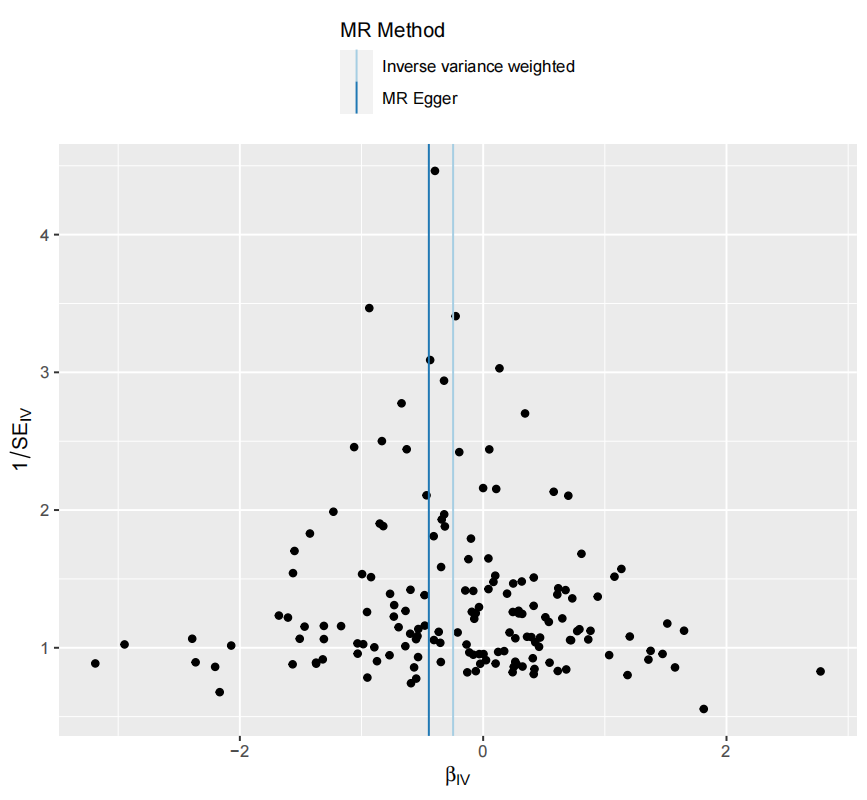

### Supplementary Figure 2. Genetic association of Sum eosinophil basophil counts with Malignant neoplasm of colon

1) Forest plot

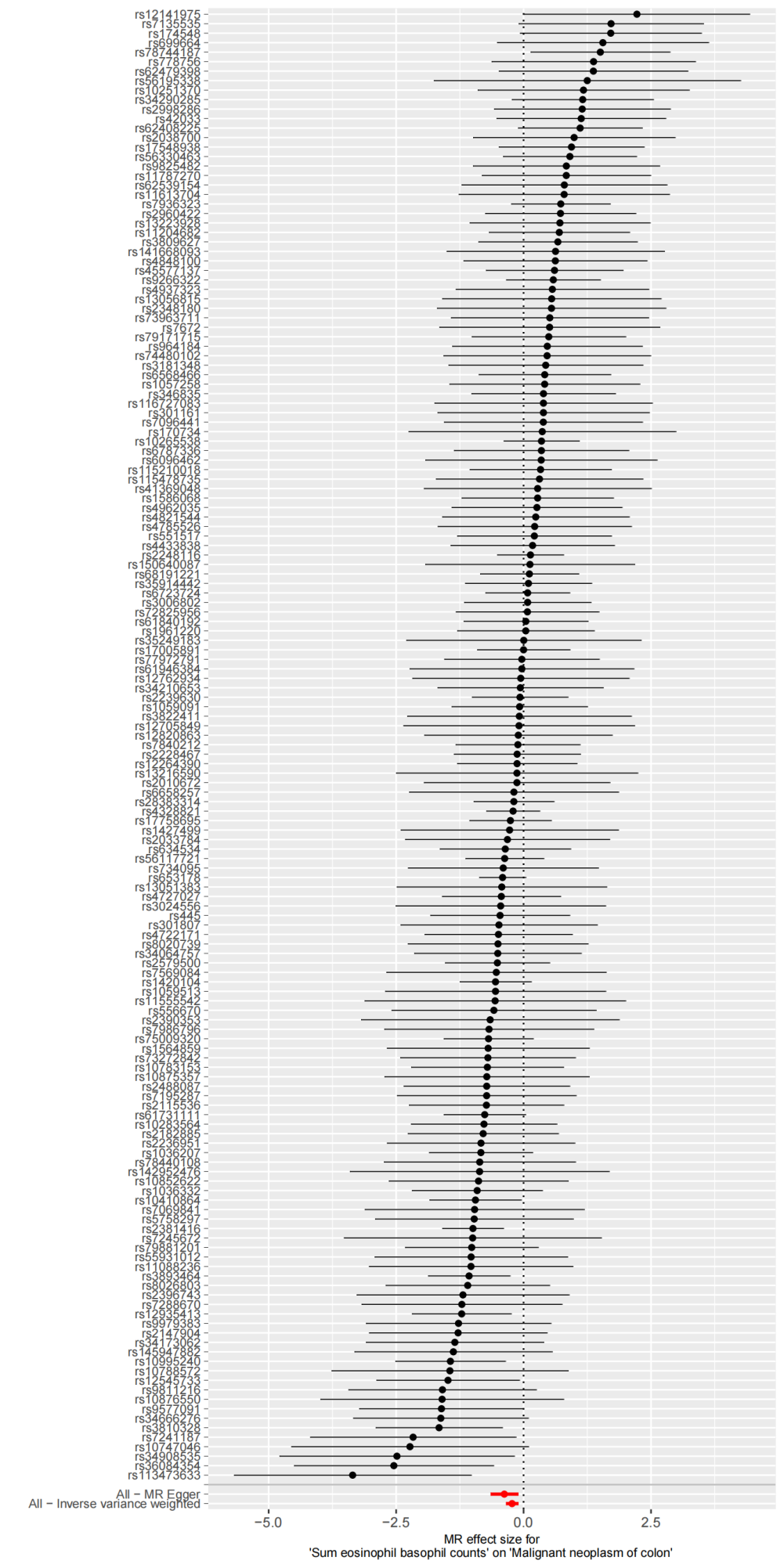

2) Leave-one-out plot

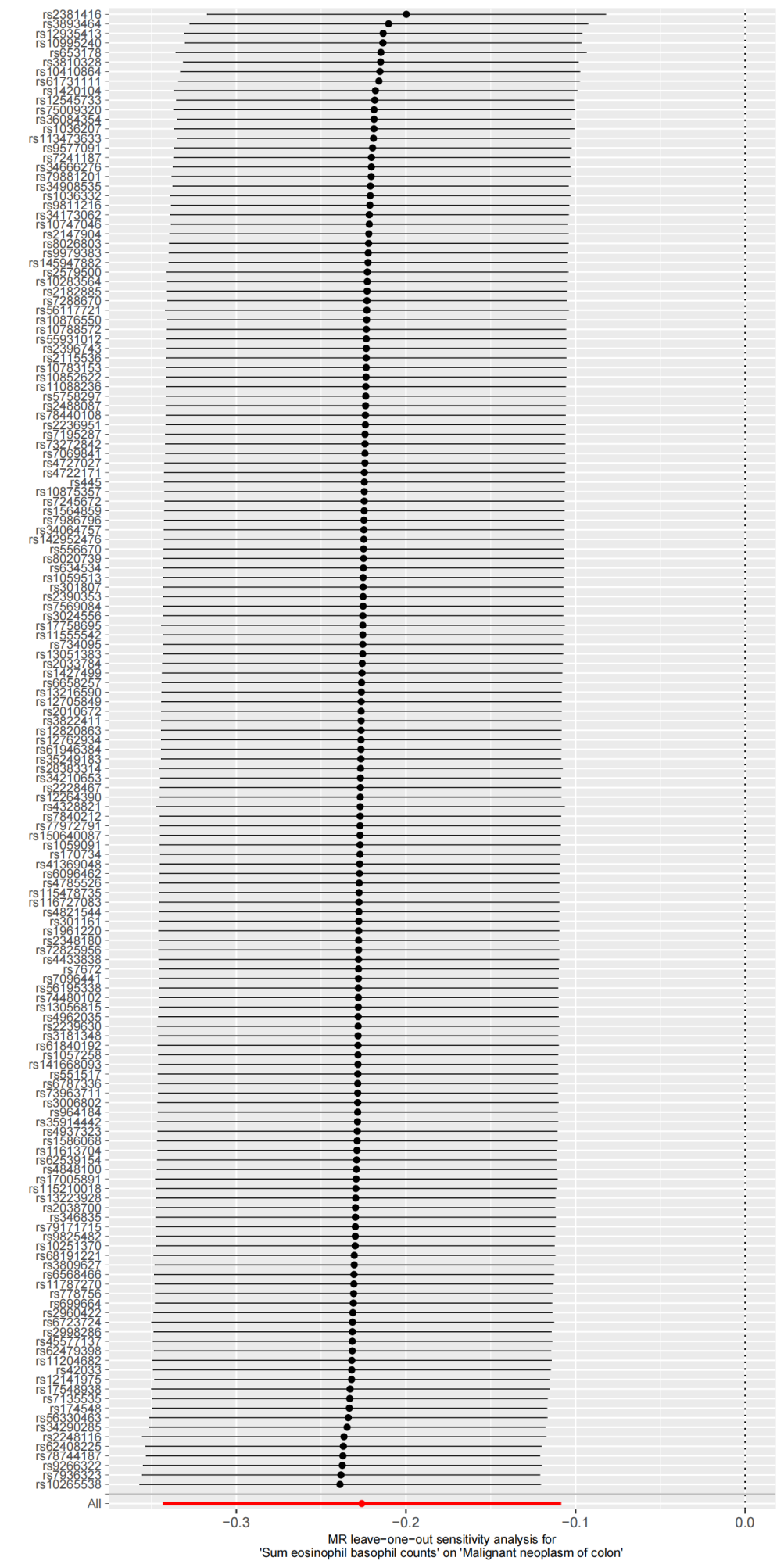

3) Scatter plot

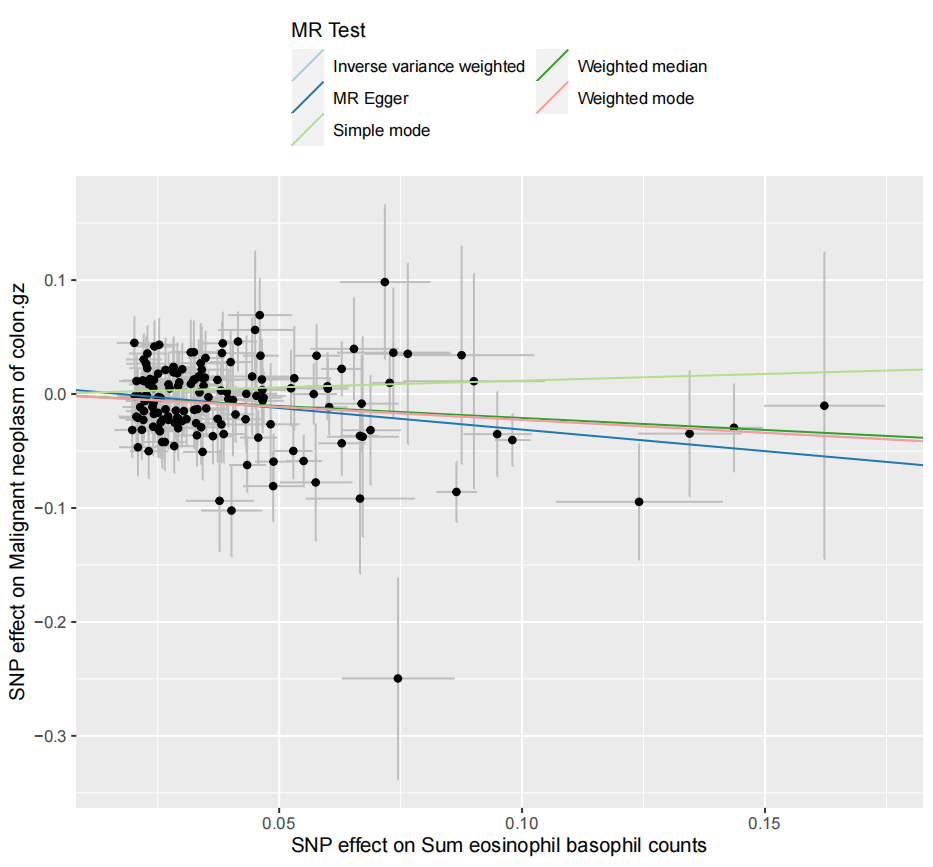

4) Funnel plot

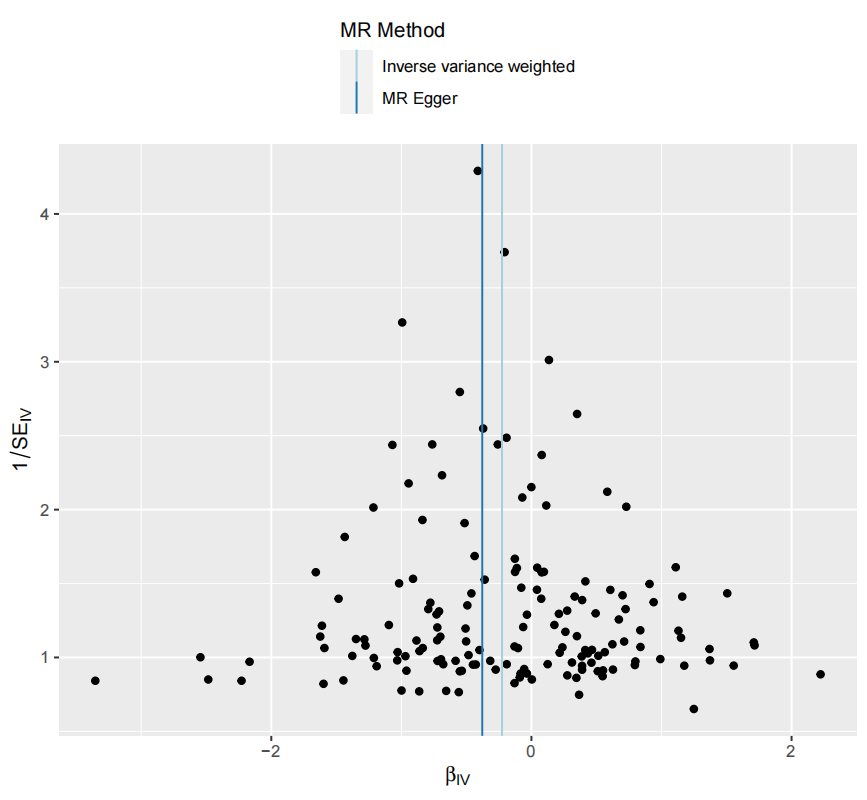

### Supplementary Figure 3. Genetic association of Hematocrit with Malignant neoplasm of ovary

1) Forest plot

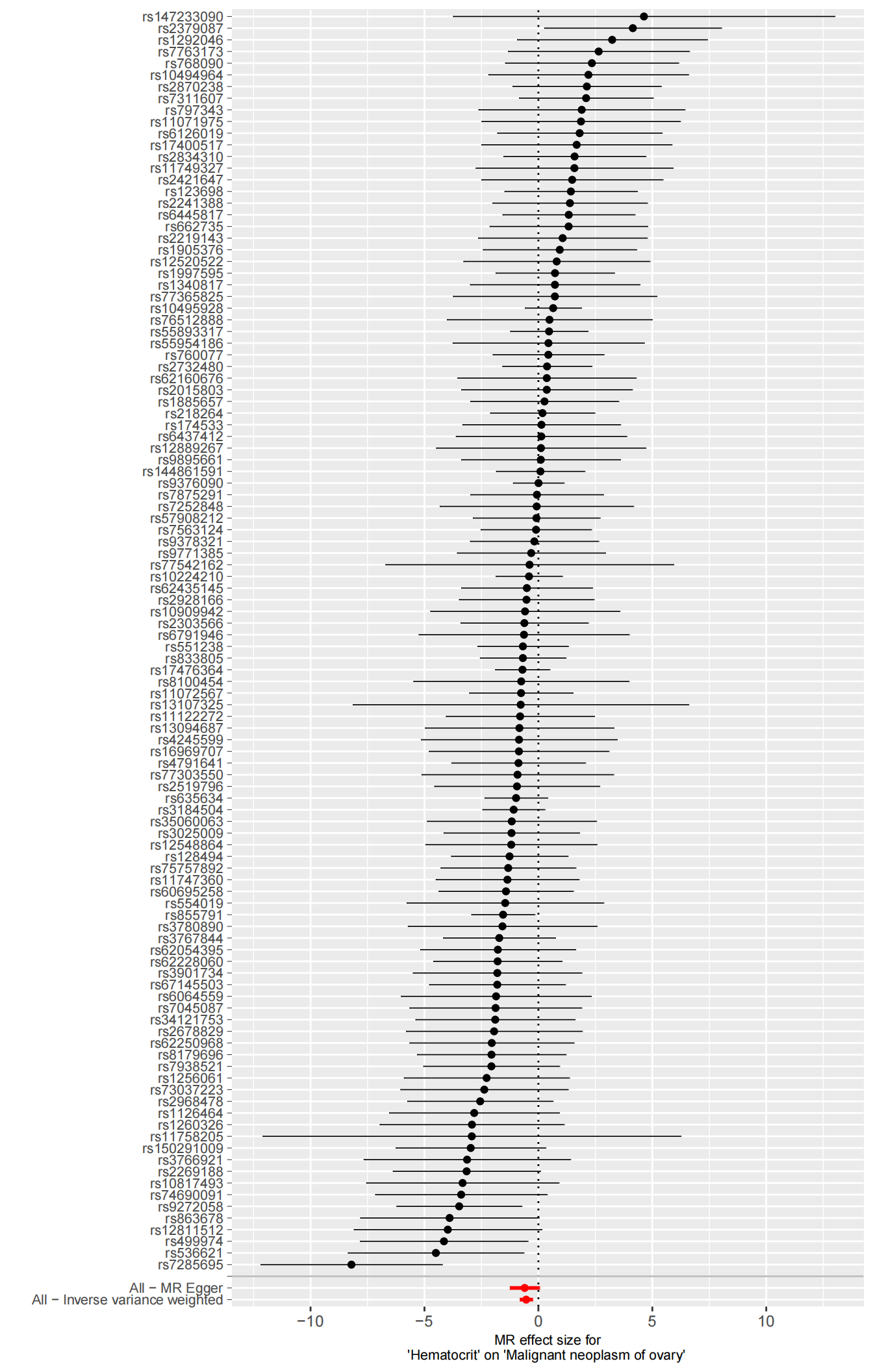

2) Leave-one-out plot

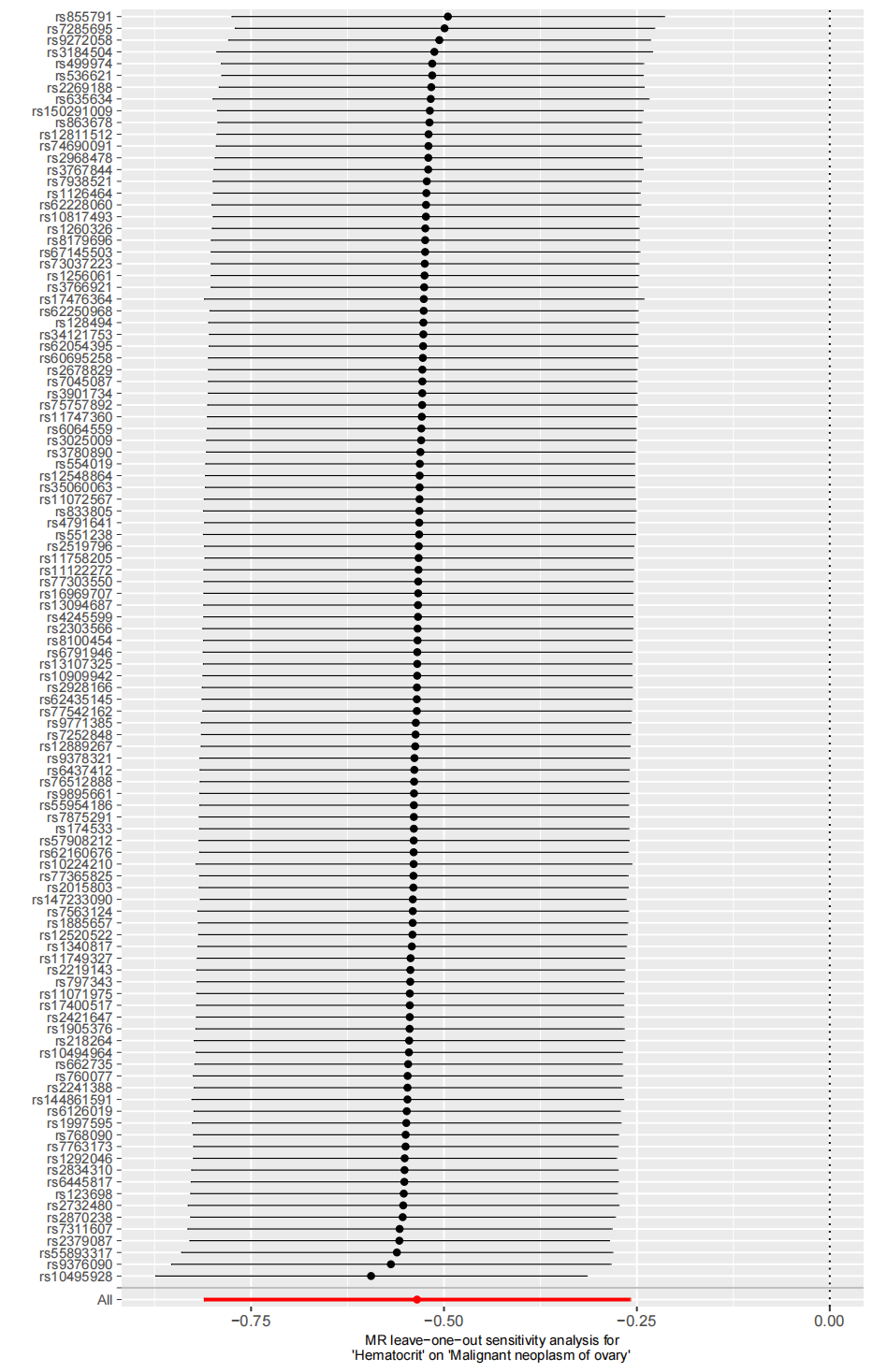

3) Scatter plot

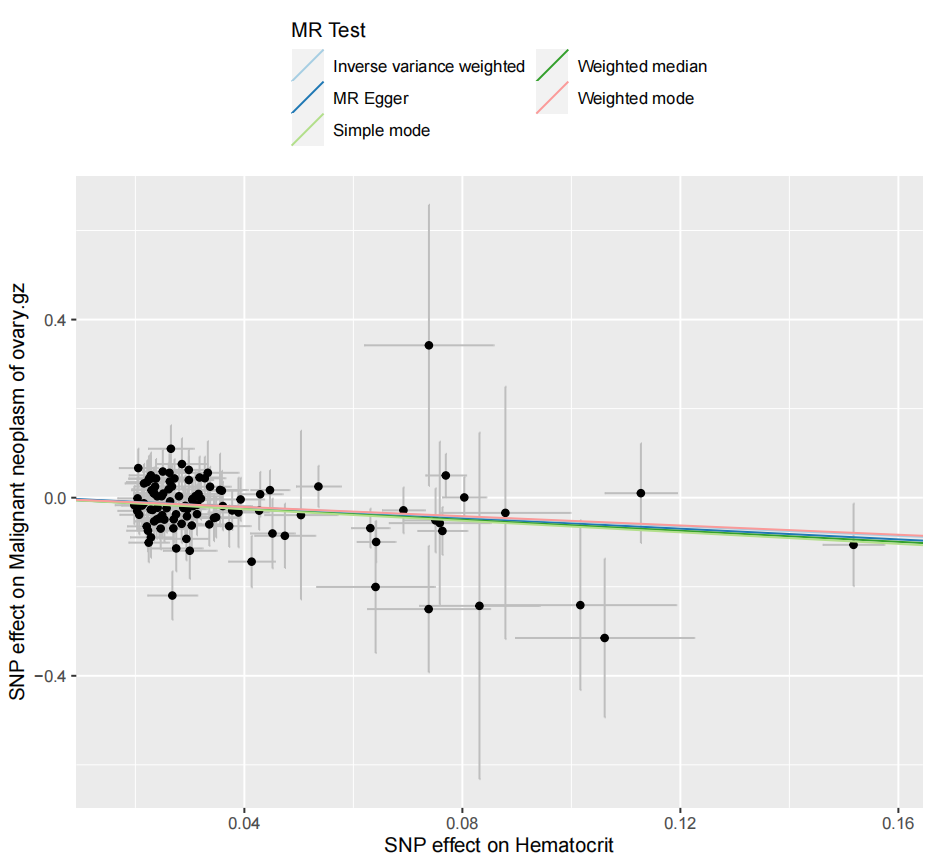

4) Funnel plot

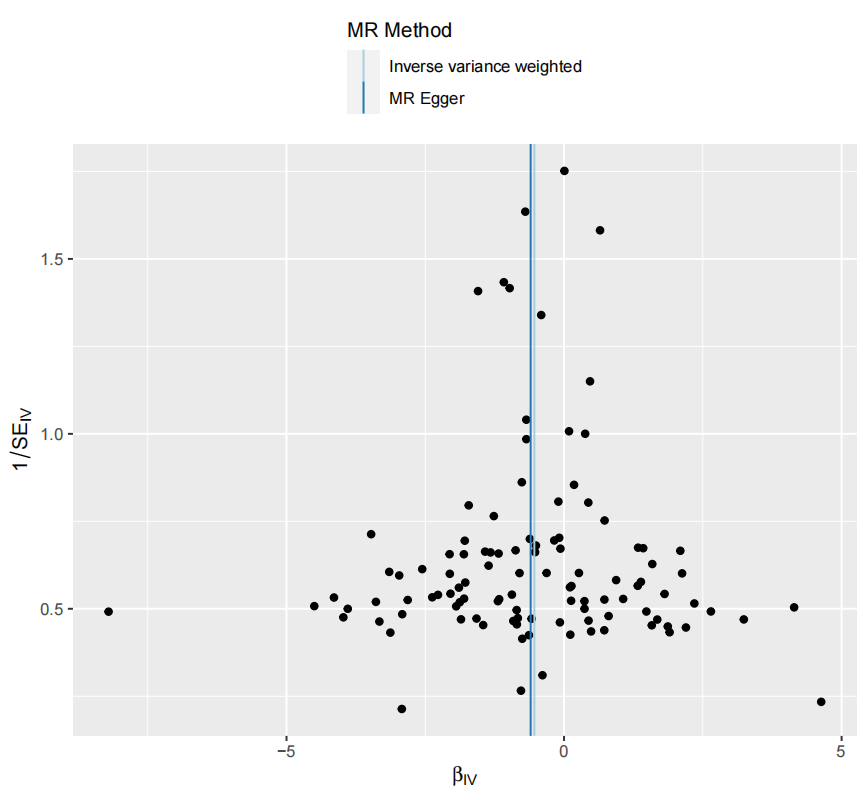

### Supplementary Figure 4. Genetic association of Eosinophil percentage of white cells with Malignant neoplasm of colon

1) Forest plot

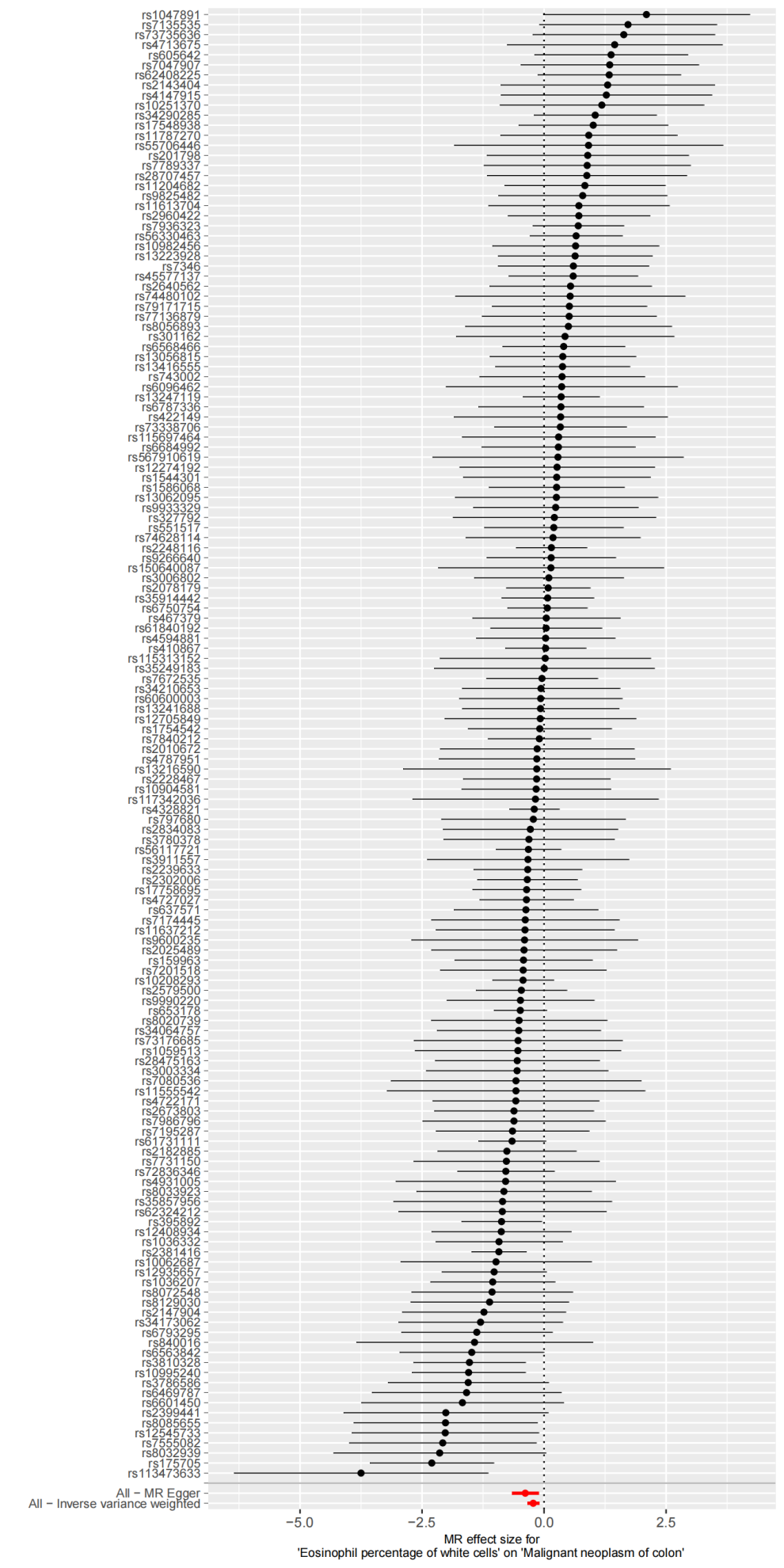

2) Leave-one-out plot

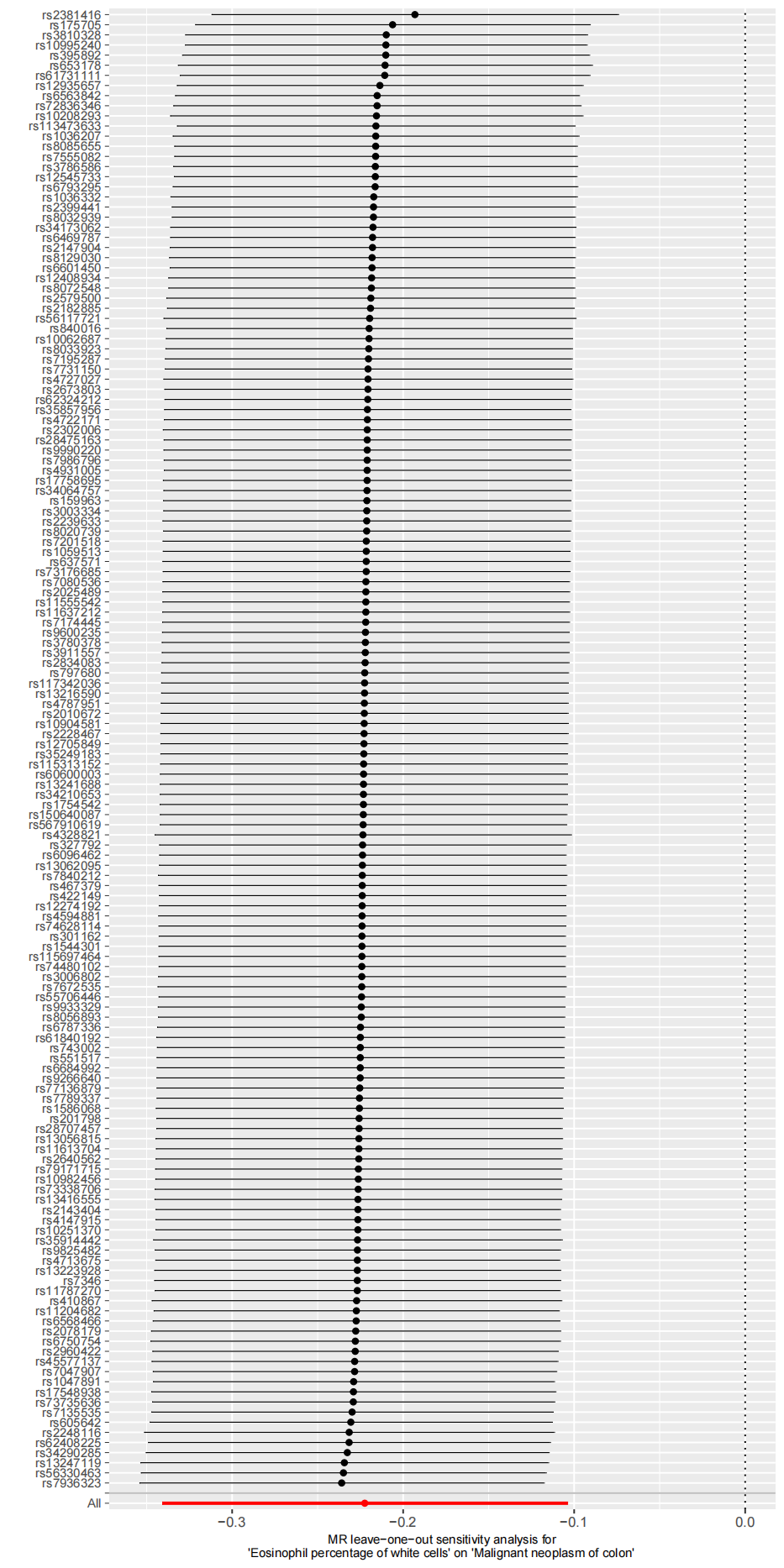

3) Scatter plot

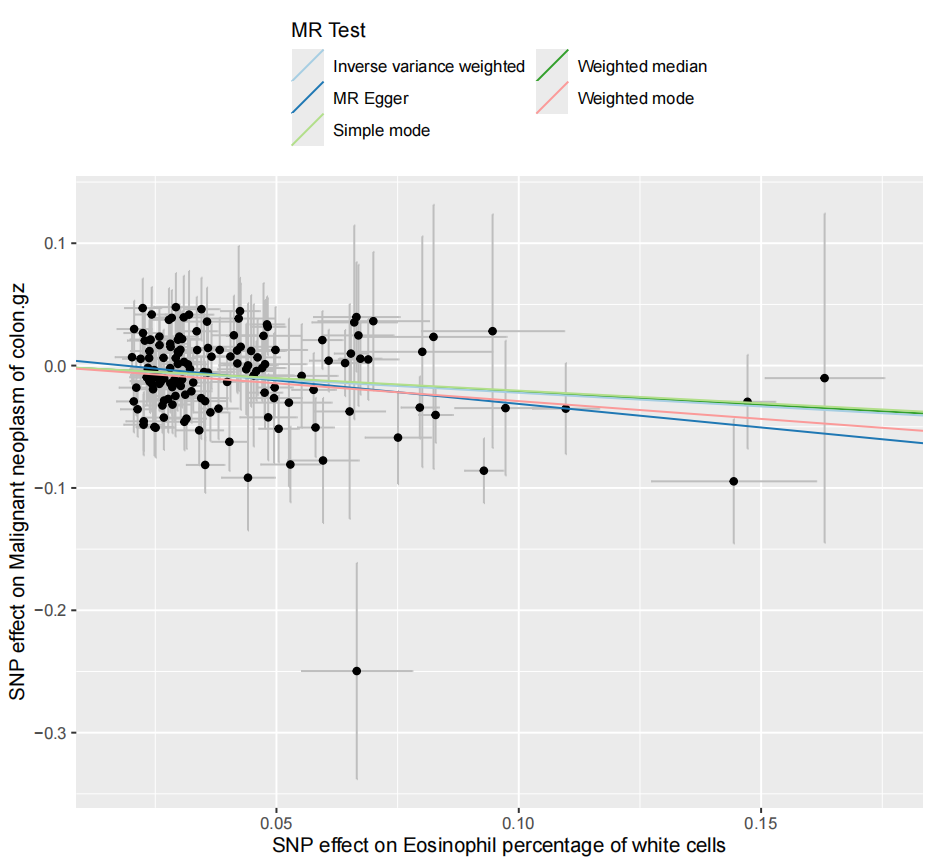

4) Funnel plot

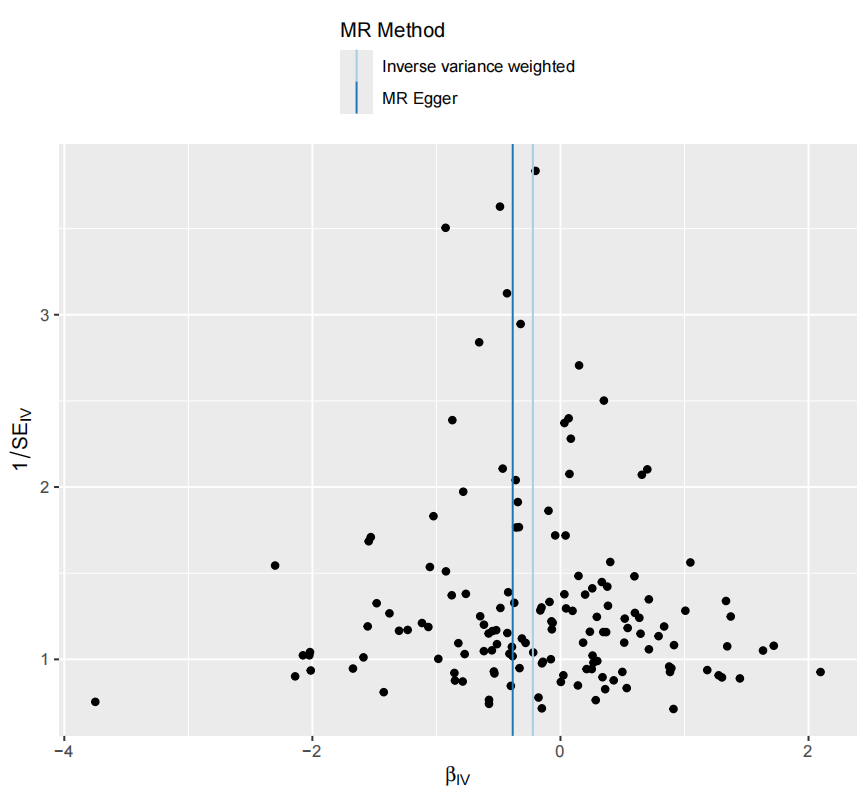

### Supplementary Figure 5. Genetic association of Sum eosinophil basophil counts with Malignant neoplasm of vulva

1) Forest plot

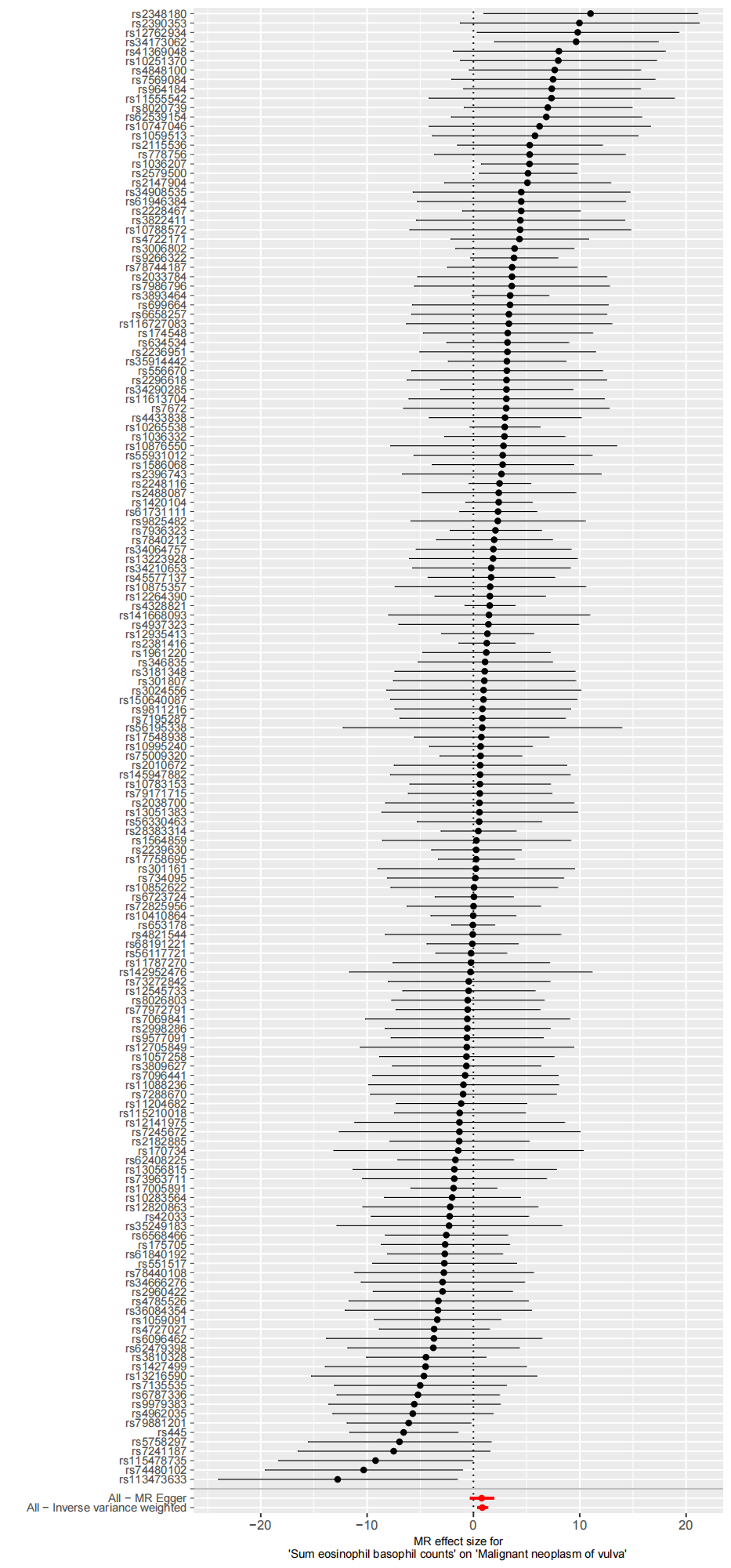

2) Leave-one-out plot

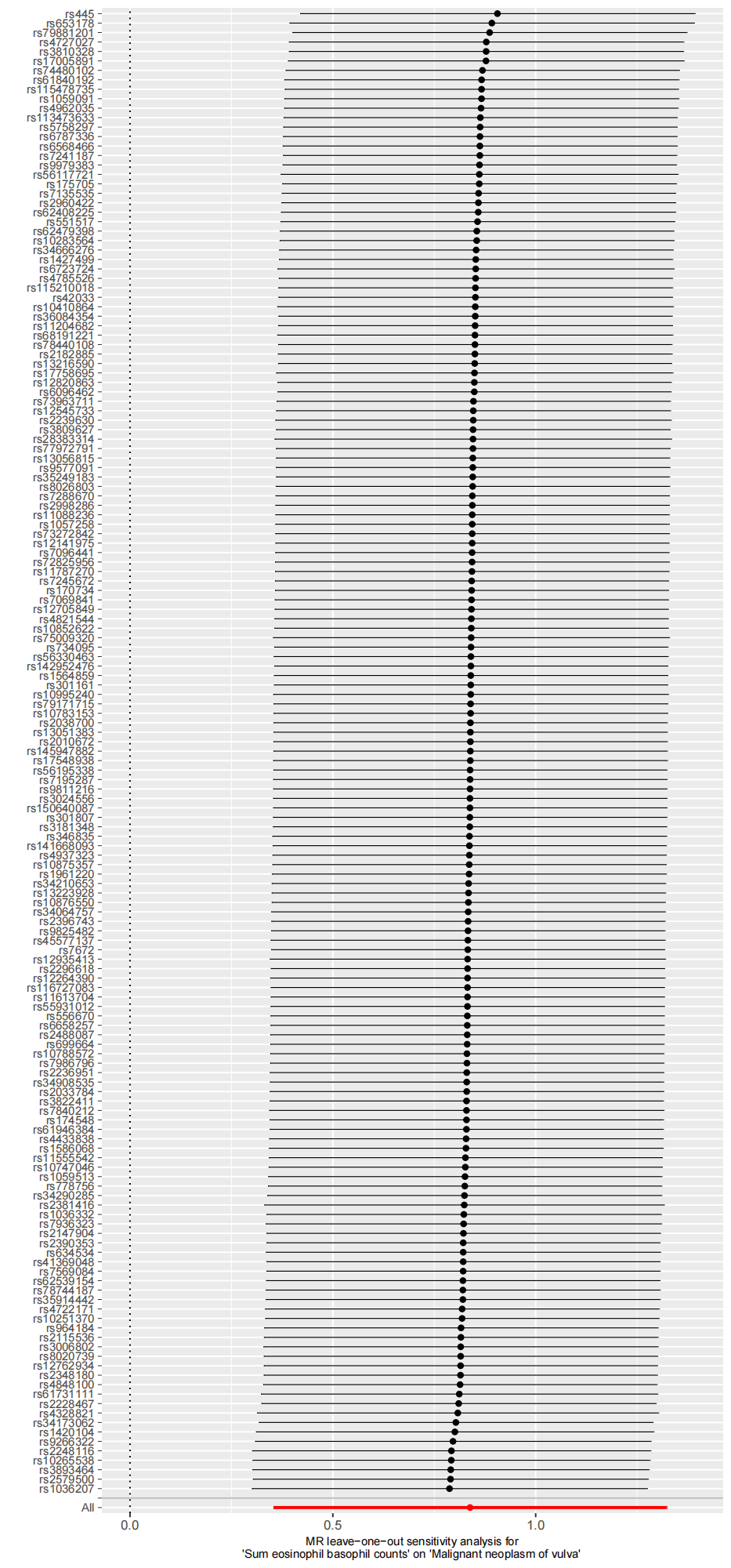

3) Scatter plot

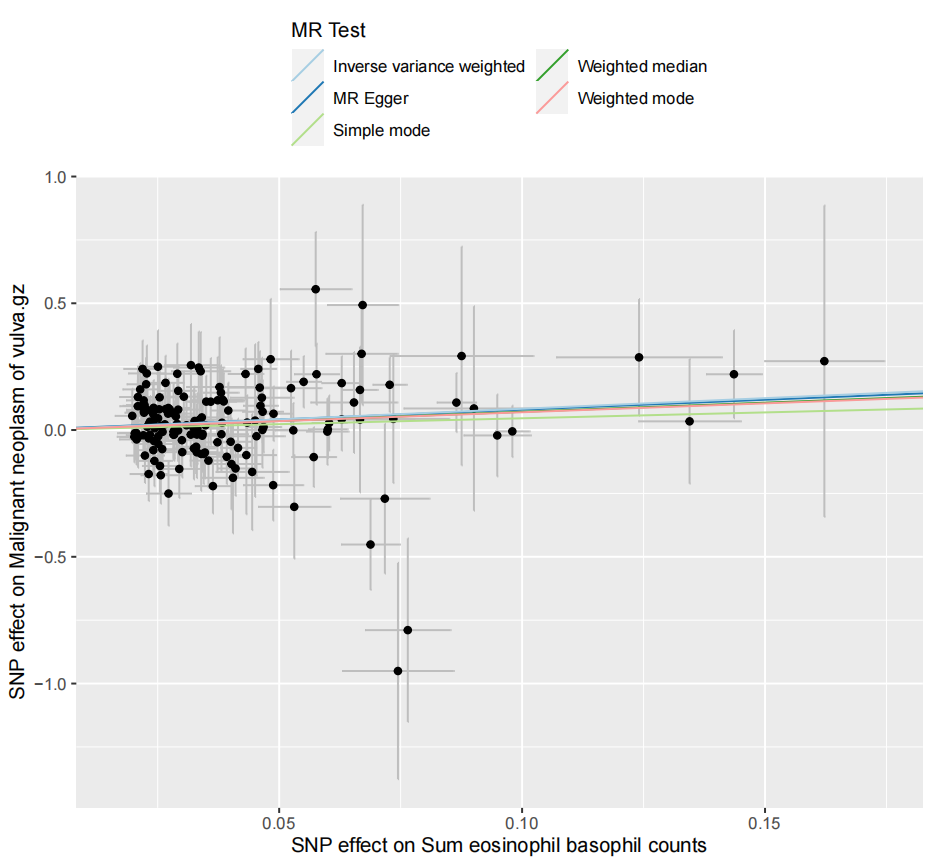

4) Funnel plot

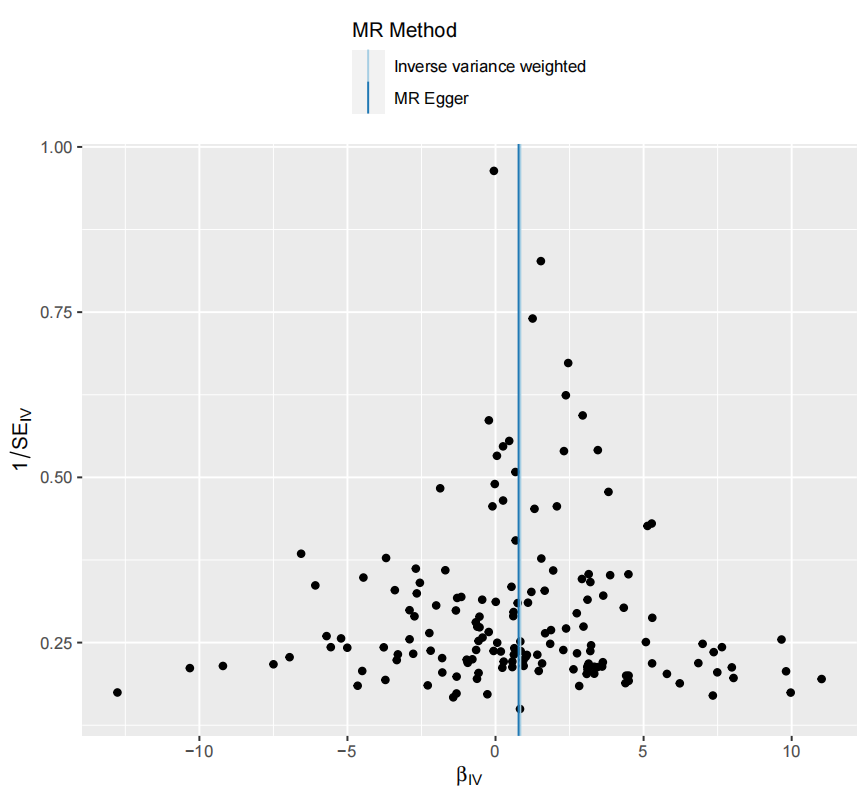

### Supplementary Figure 6. Genetic association of Eosinophil percentage of granulocytes with Malignant neoplasm of colon

1) Forest plot

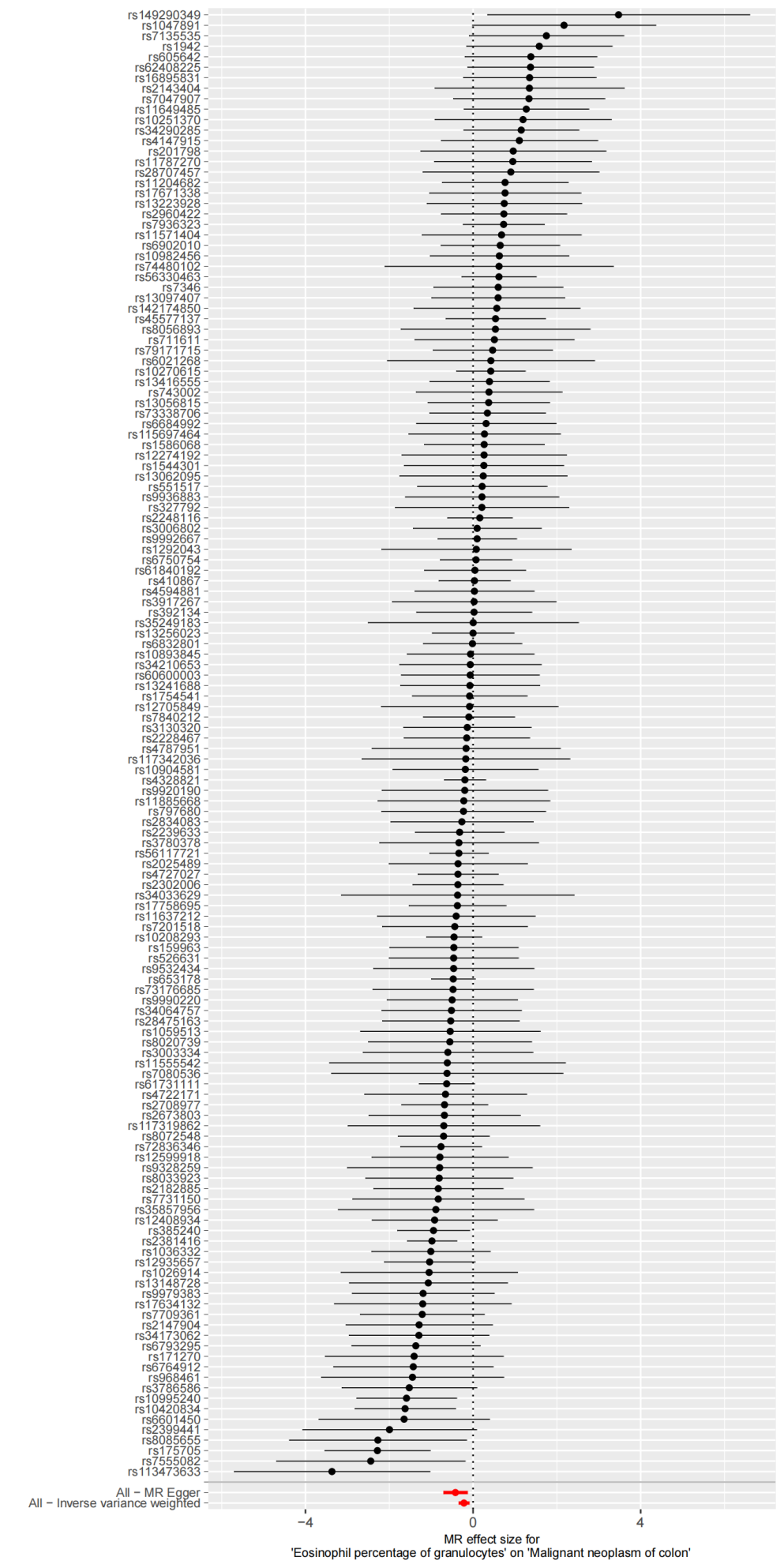

2) Leave-one-out plot

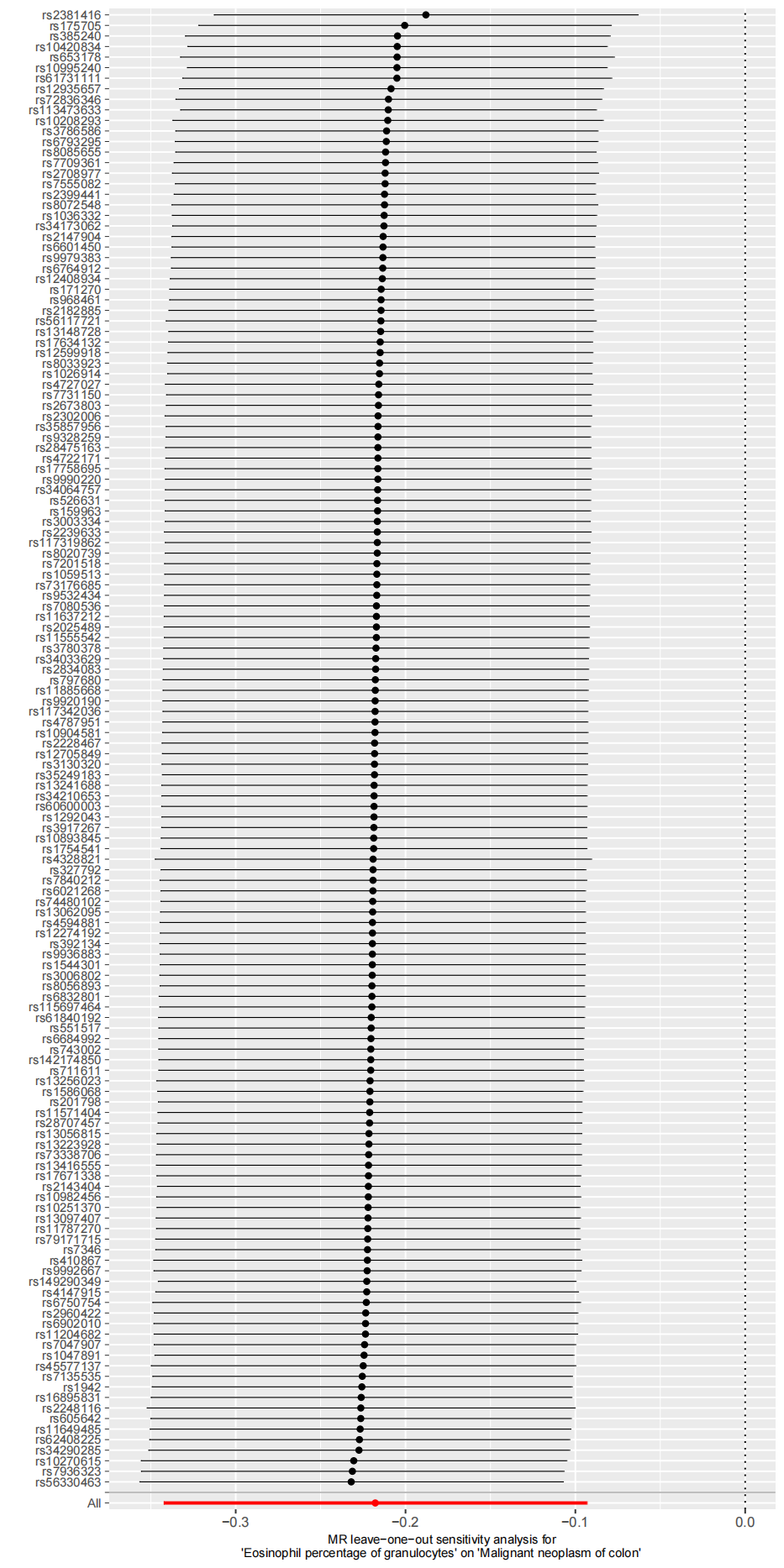

3) Scatter plot

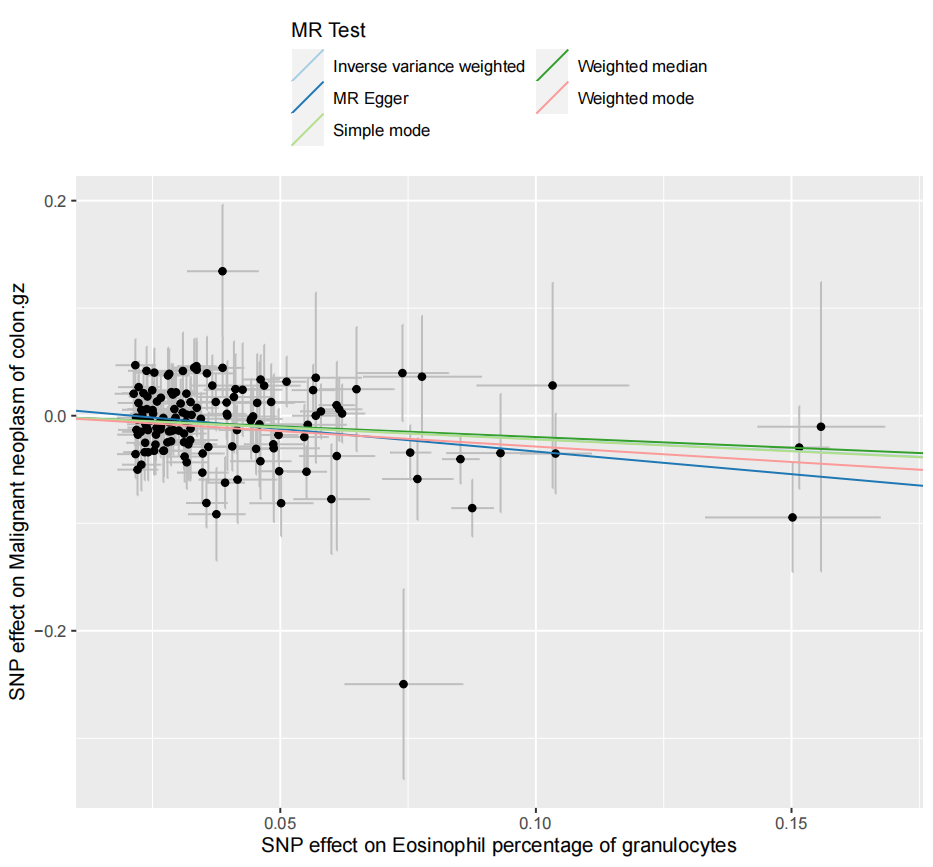

4) Funnel plot

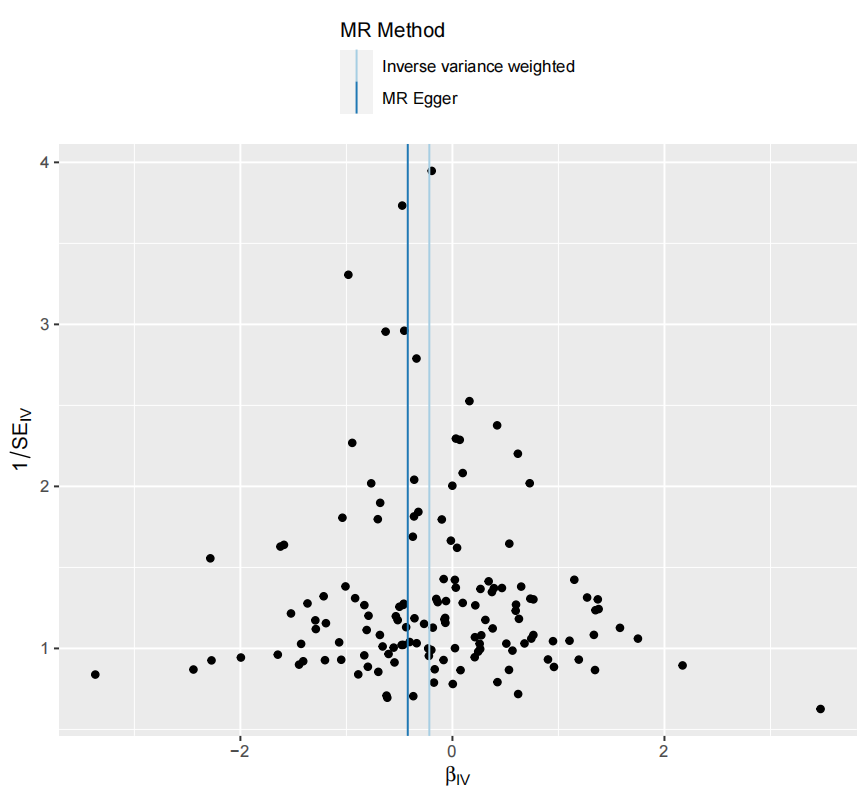

### Supplementary Figure 7. Genetic association of Plateletcrit with Malignant cancer of tonsil and base of tongue

1) Forest plot

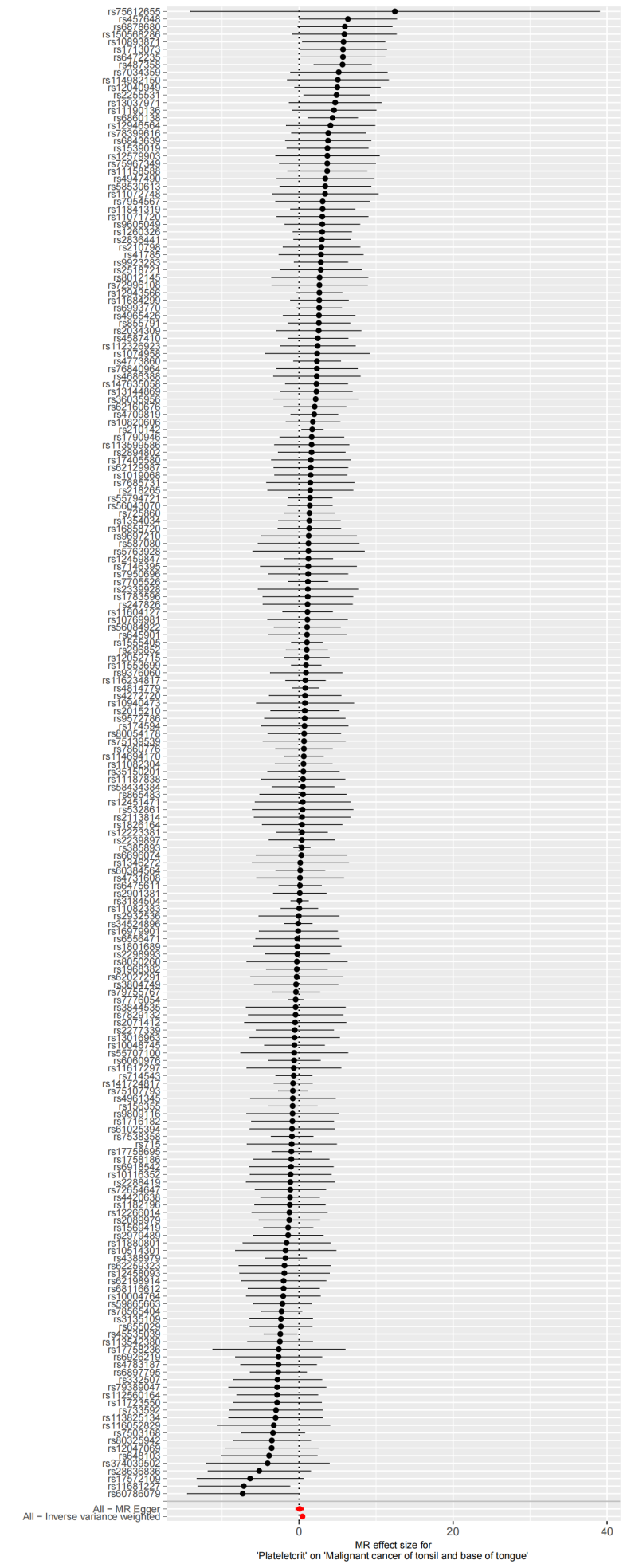

2)Leave-one-out plot

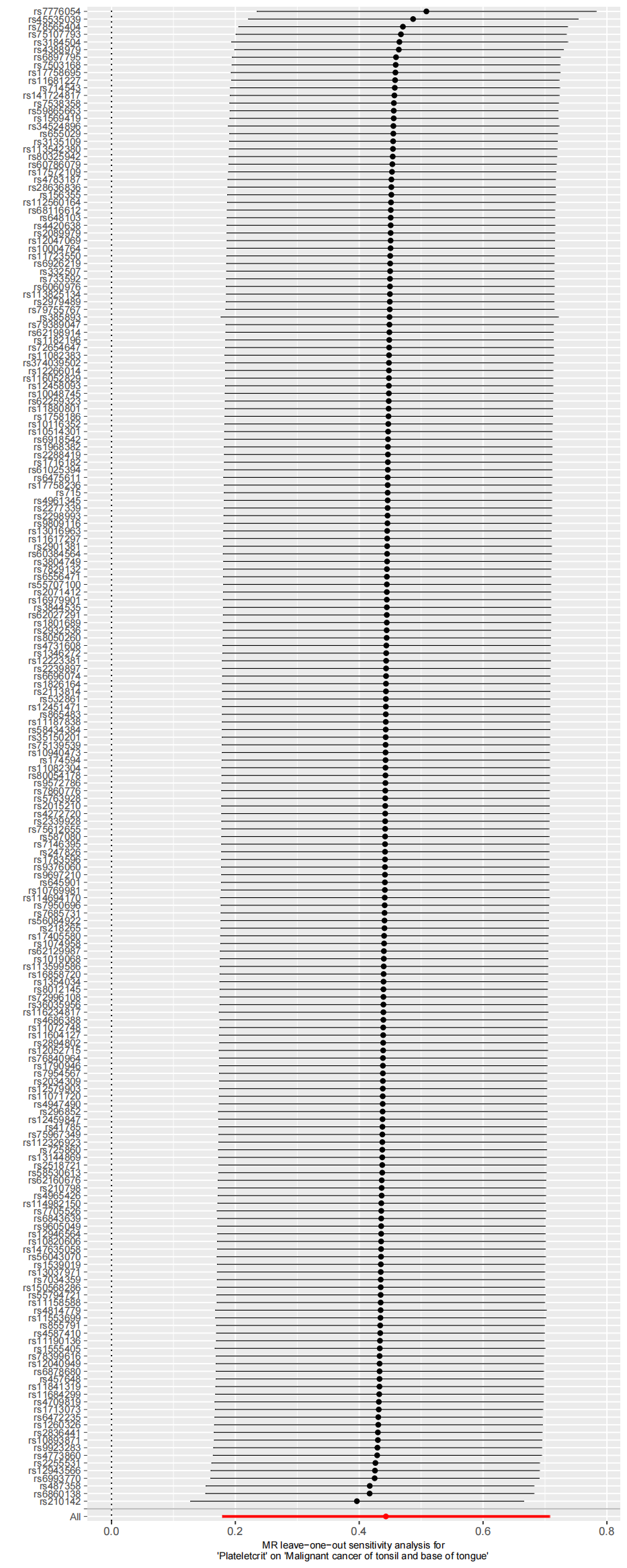

3)Scatter plot

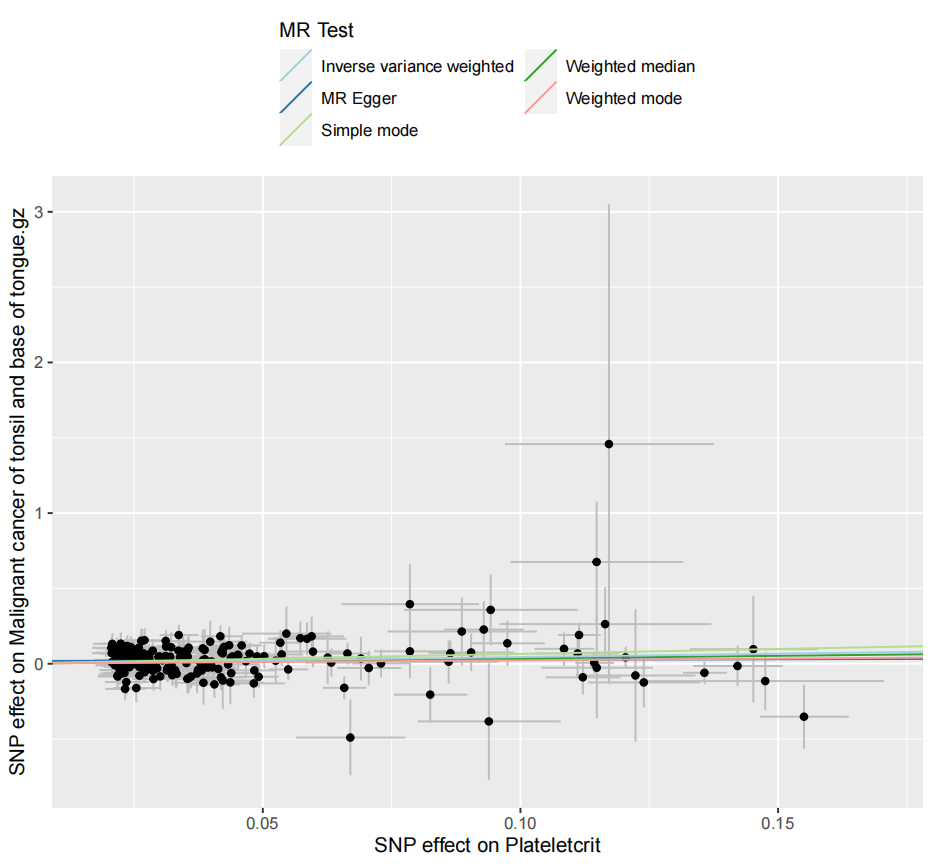

4) Funnel plot

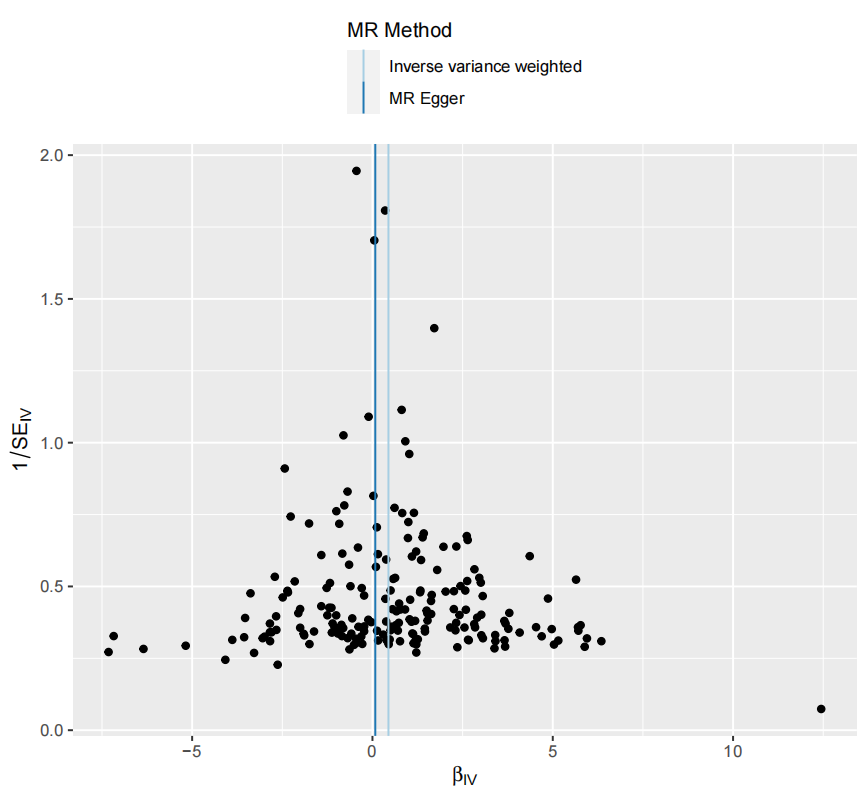

### Supplementary Figure 8. Genetic association of High light scatter reticulocyte percentage of red cells with Malignant neoplasm of stomach

1) Forest plot

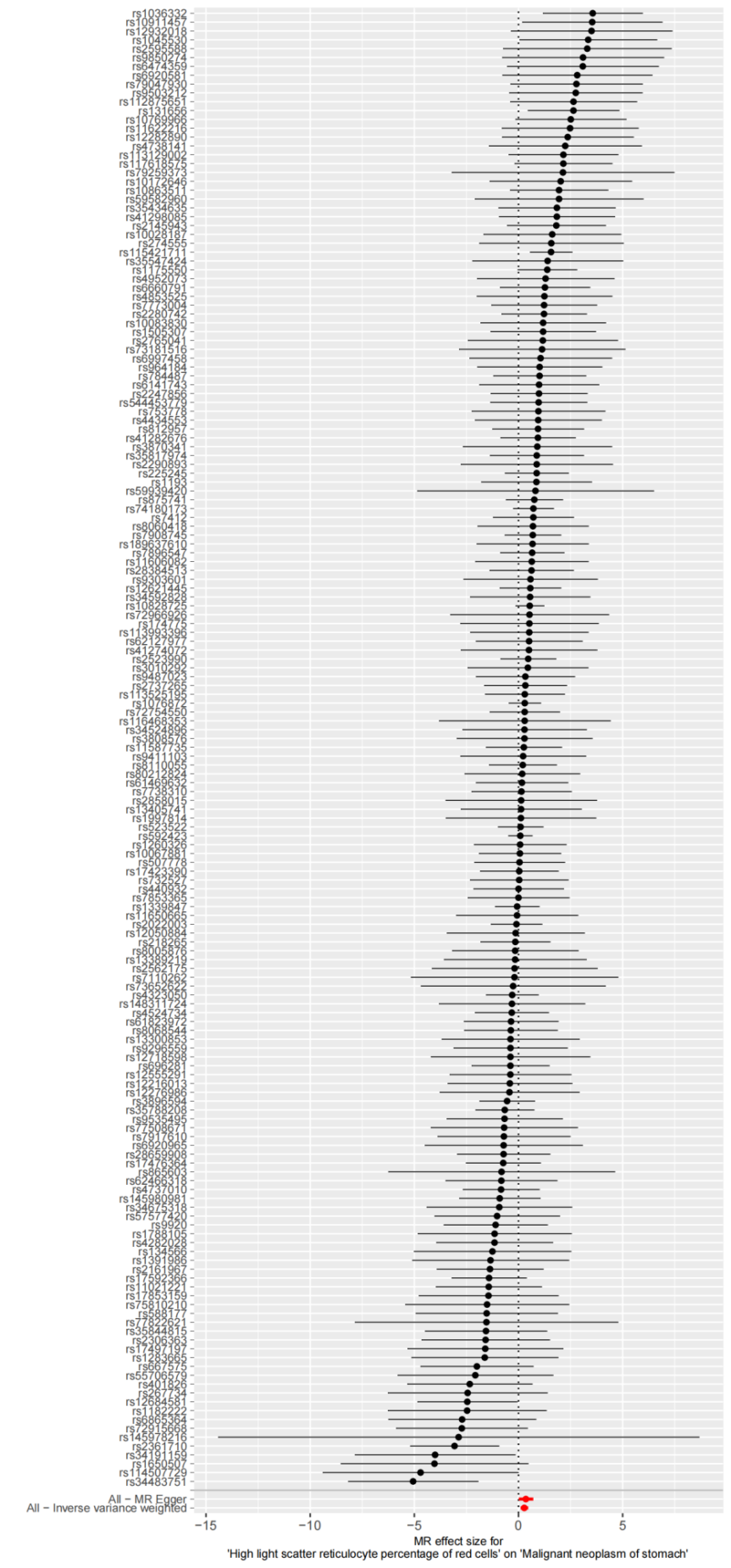

2)Leave-one-out plot

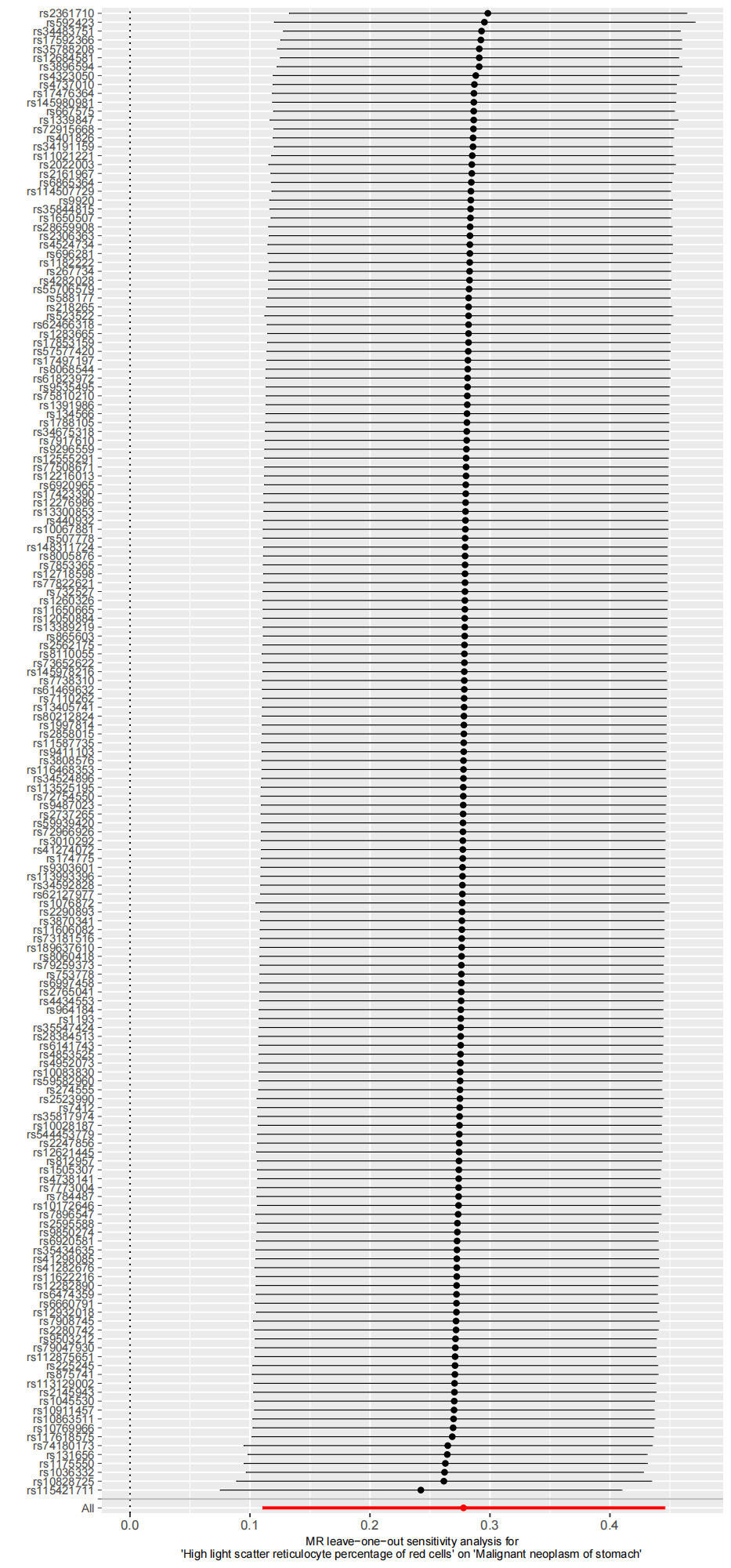

3)Scatter plot

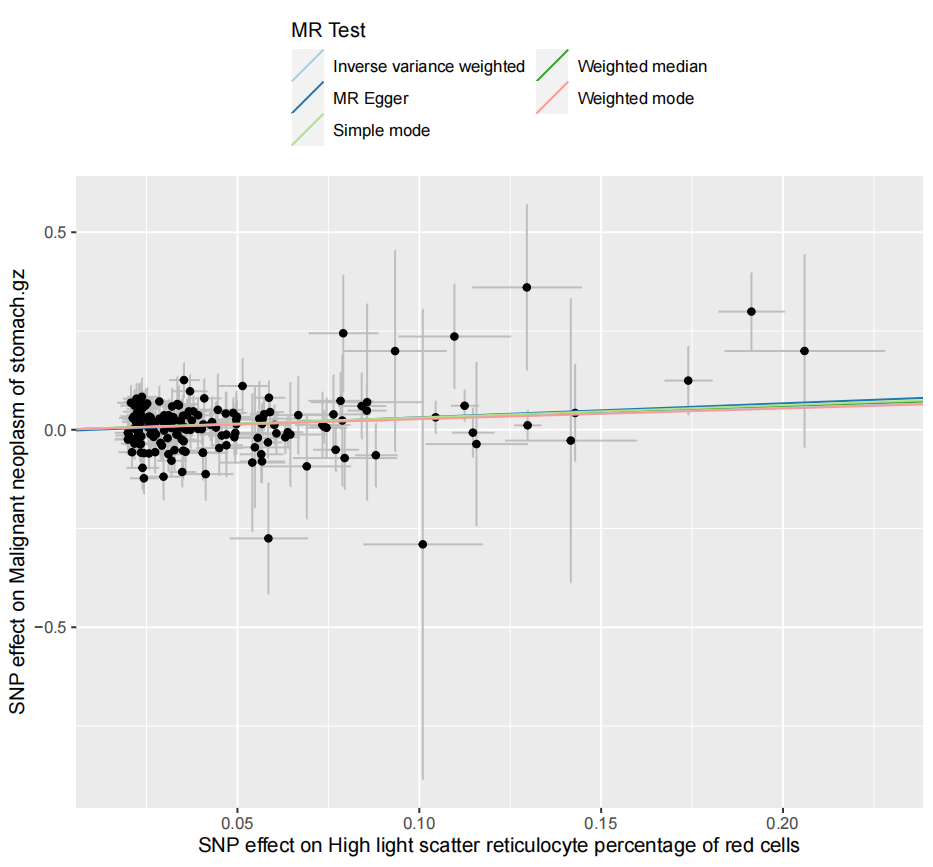

4) Funnel plot

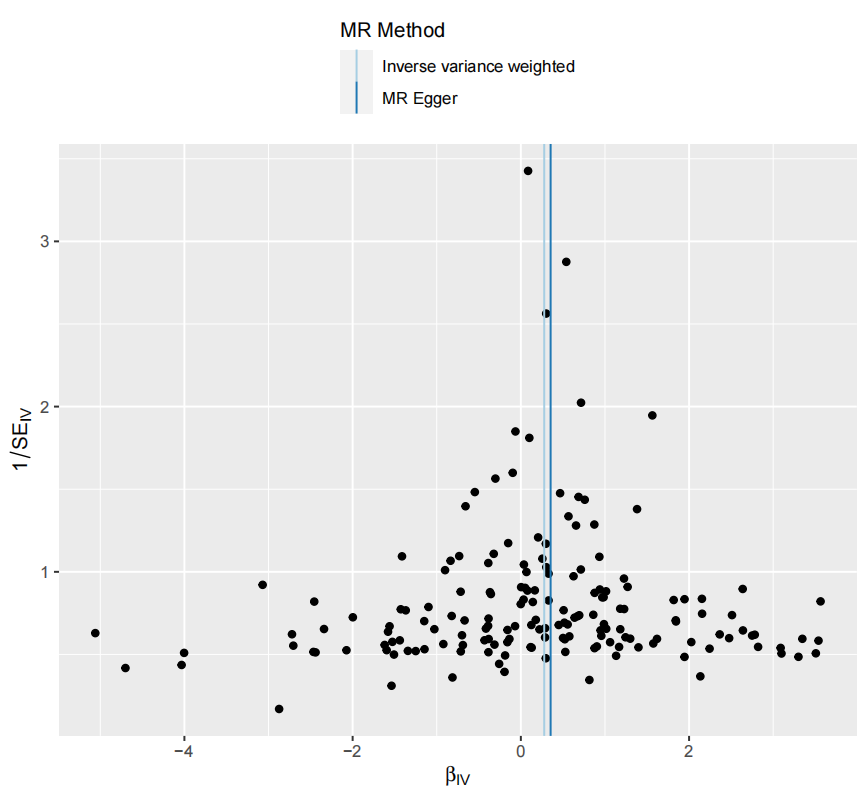

### Supplementary Figure 9. Genetic association of Eosinophil percentage of granulocytes with Malignant melanoma

1) Forest plot

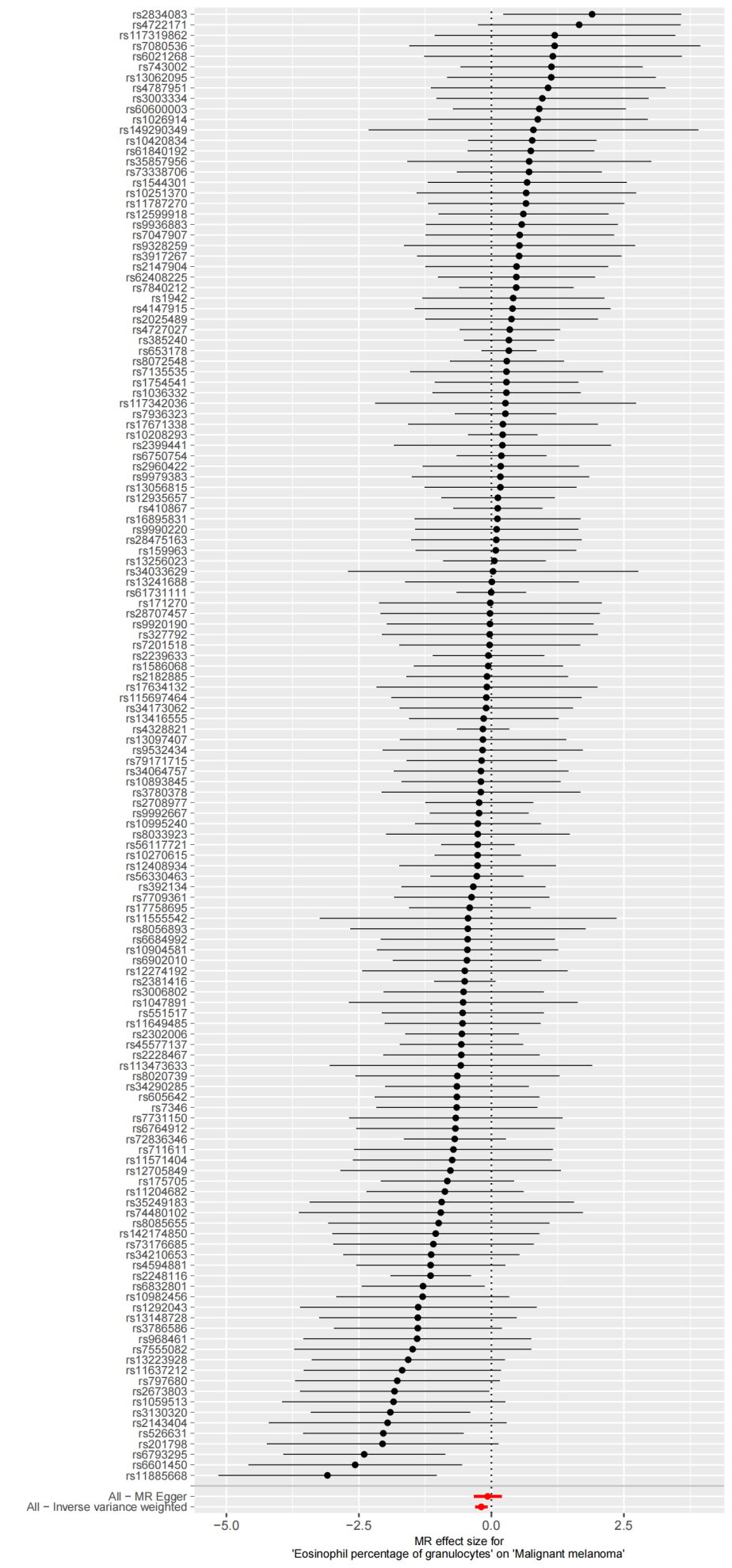

2) Leave-one-out plot

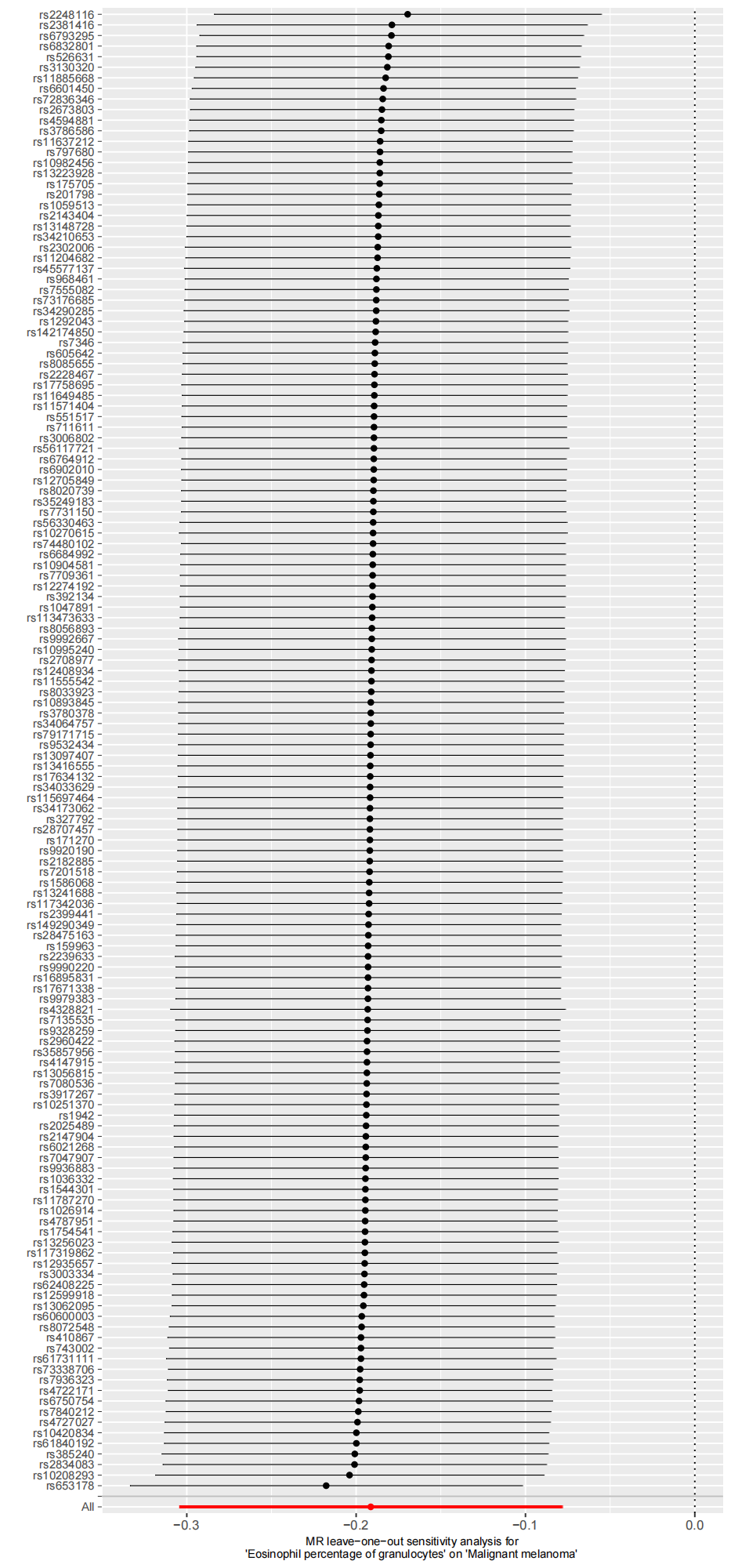

3) Scatter plot

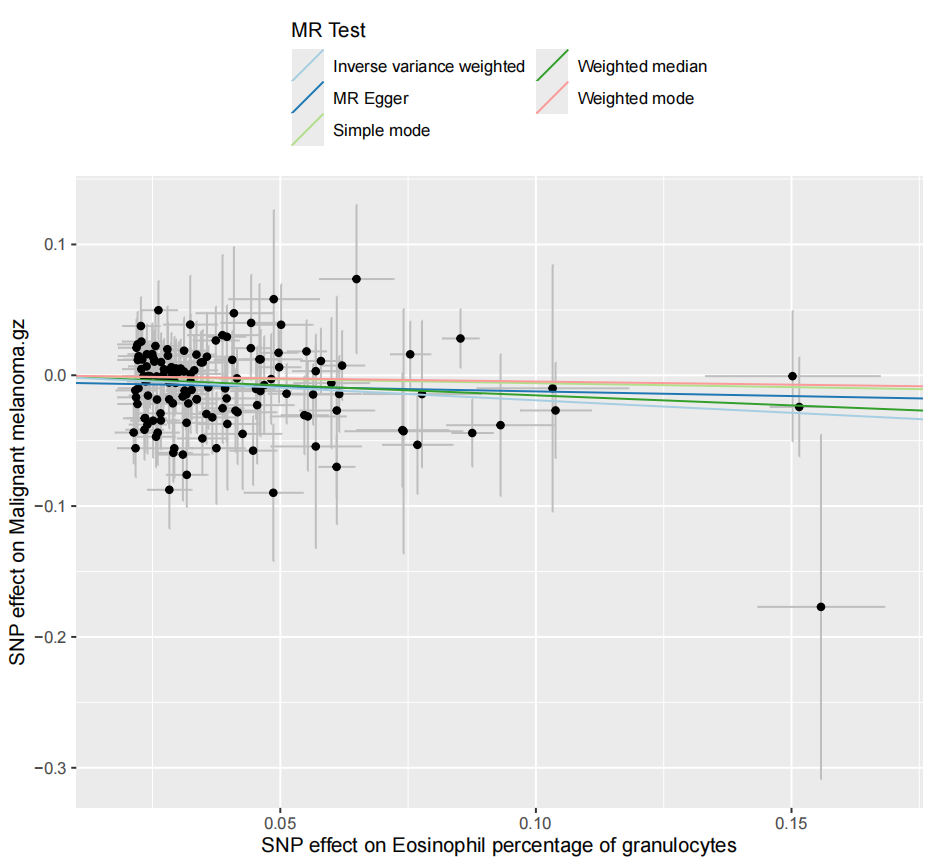

4) Funnel plot

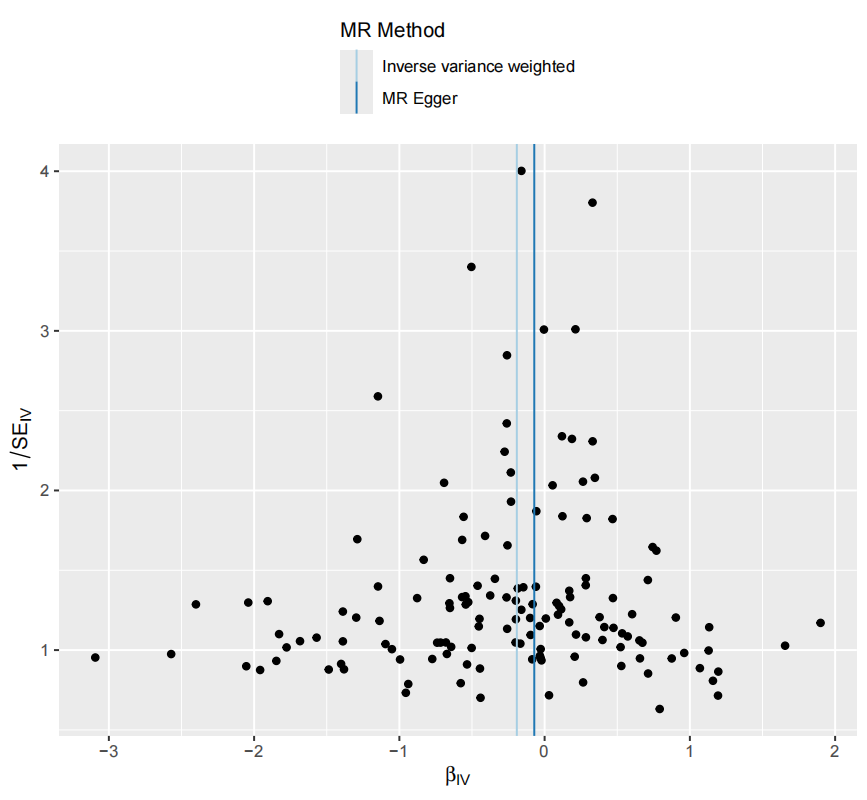

### Supplementary Figure 10. Genetic association of Red blood cell count with Malignant neoplasm of head and neck

1) Forest plot

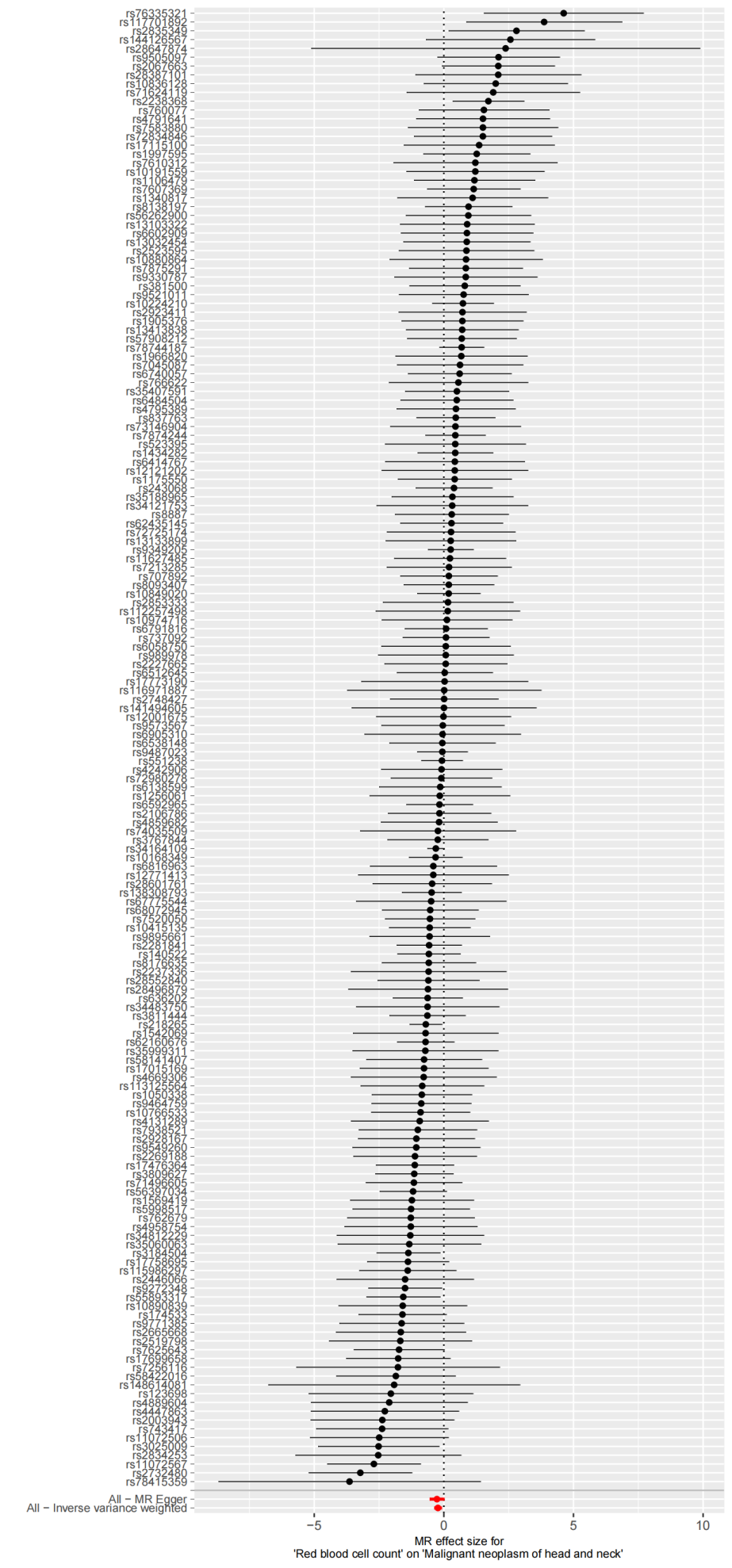

2) Leave-one-out plot

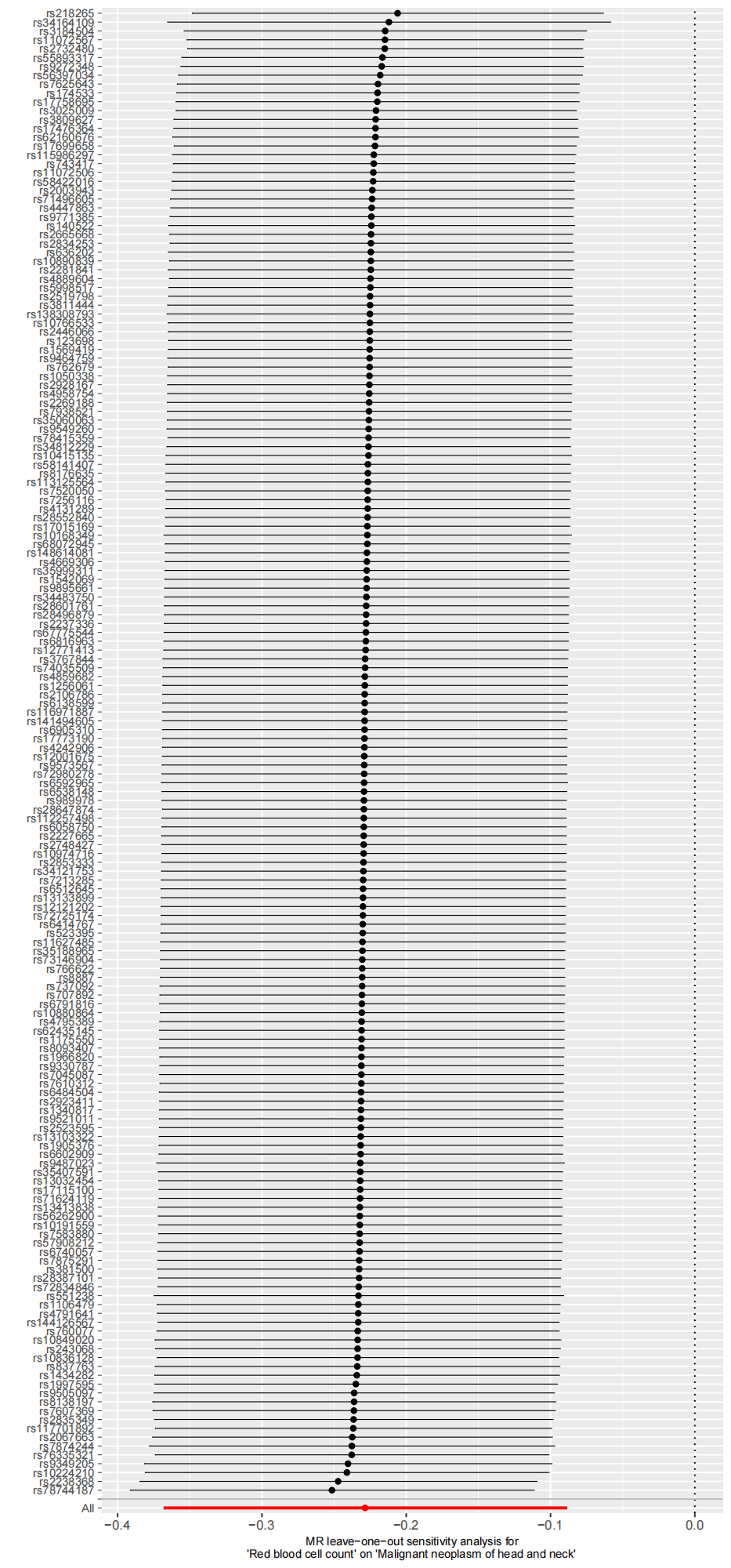

3) Scatter plot

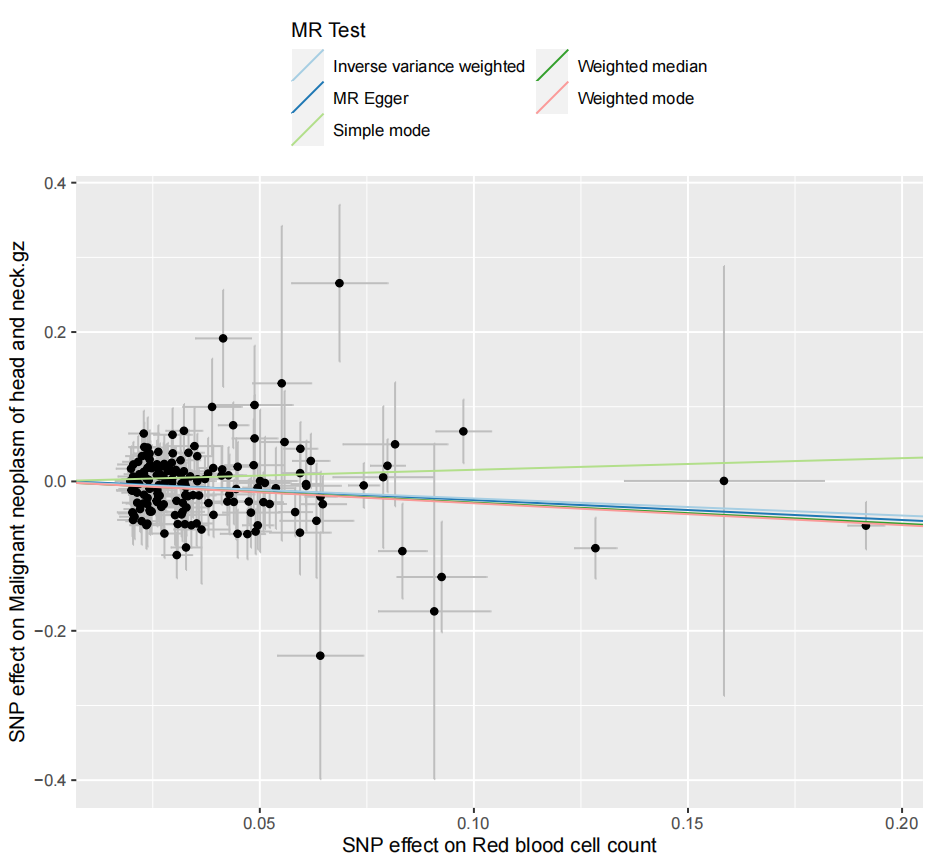

4) Funnel plot

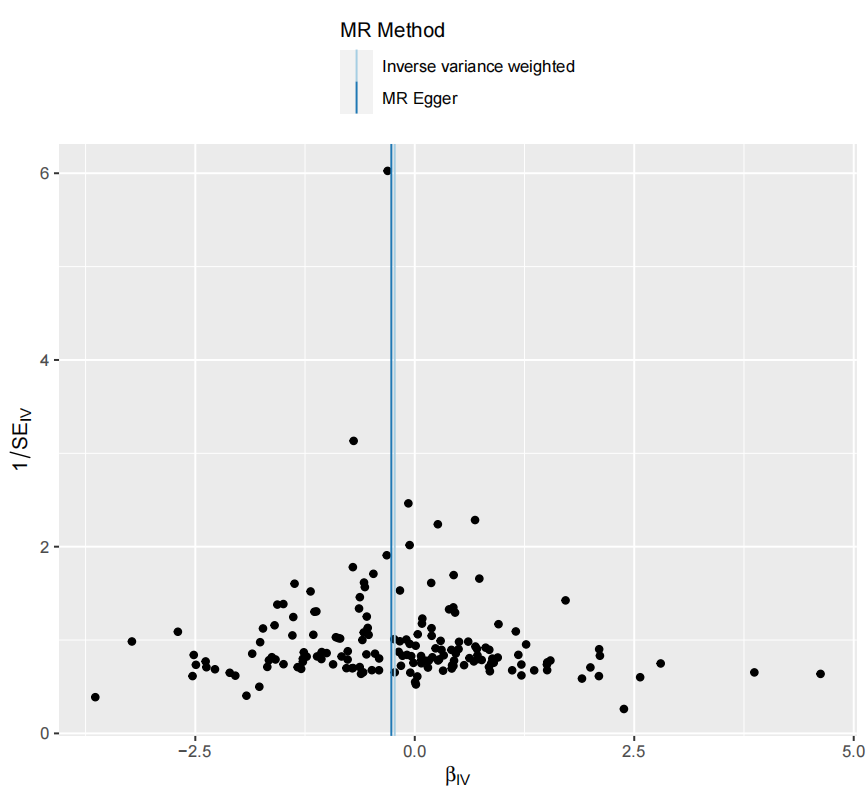

### Supplementary Figure 11. Genetic association of Red cell distribution width with Malignant neoplasm of urinary organs

1) Forest plot

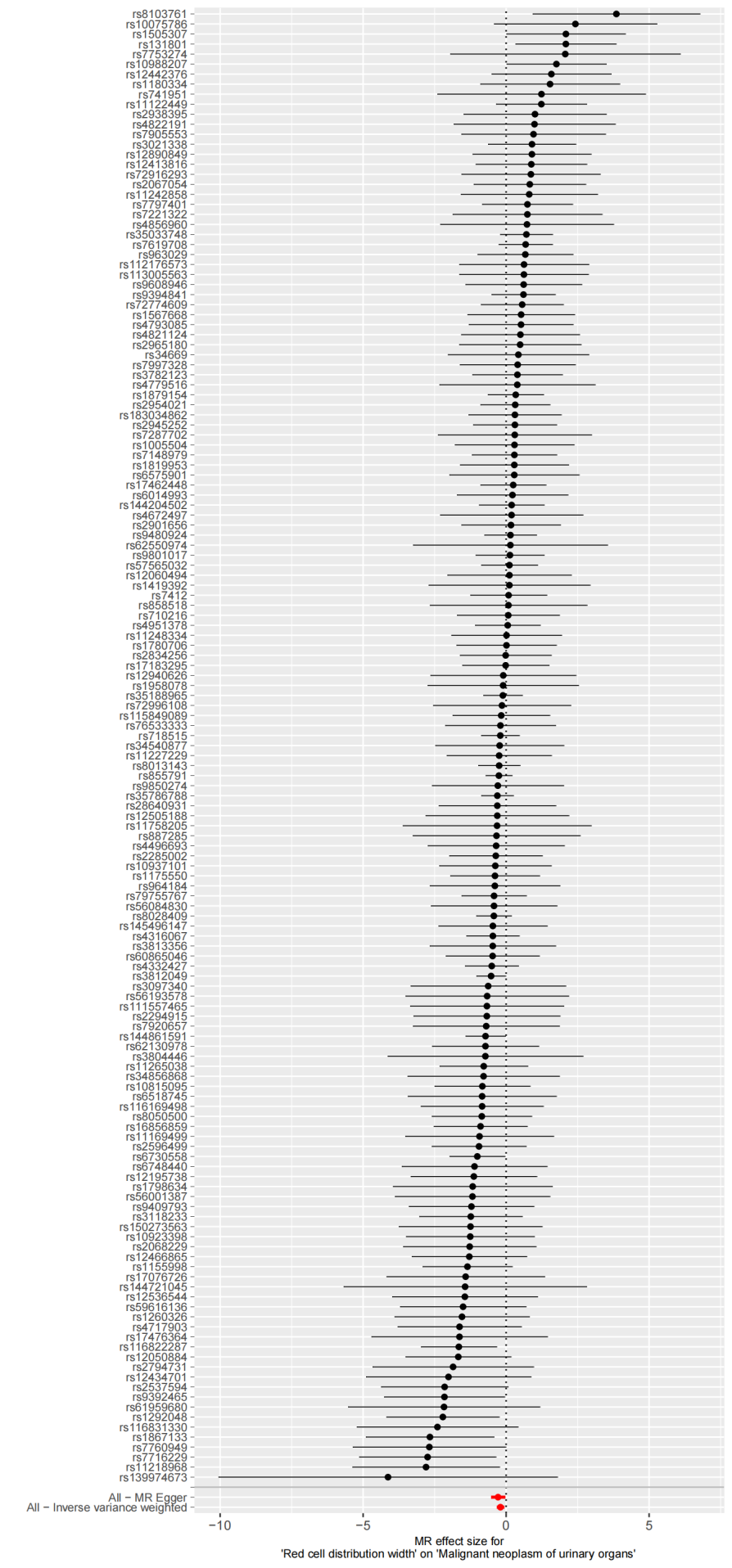

2) Leave-one-out plot

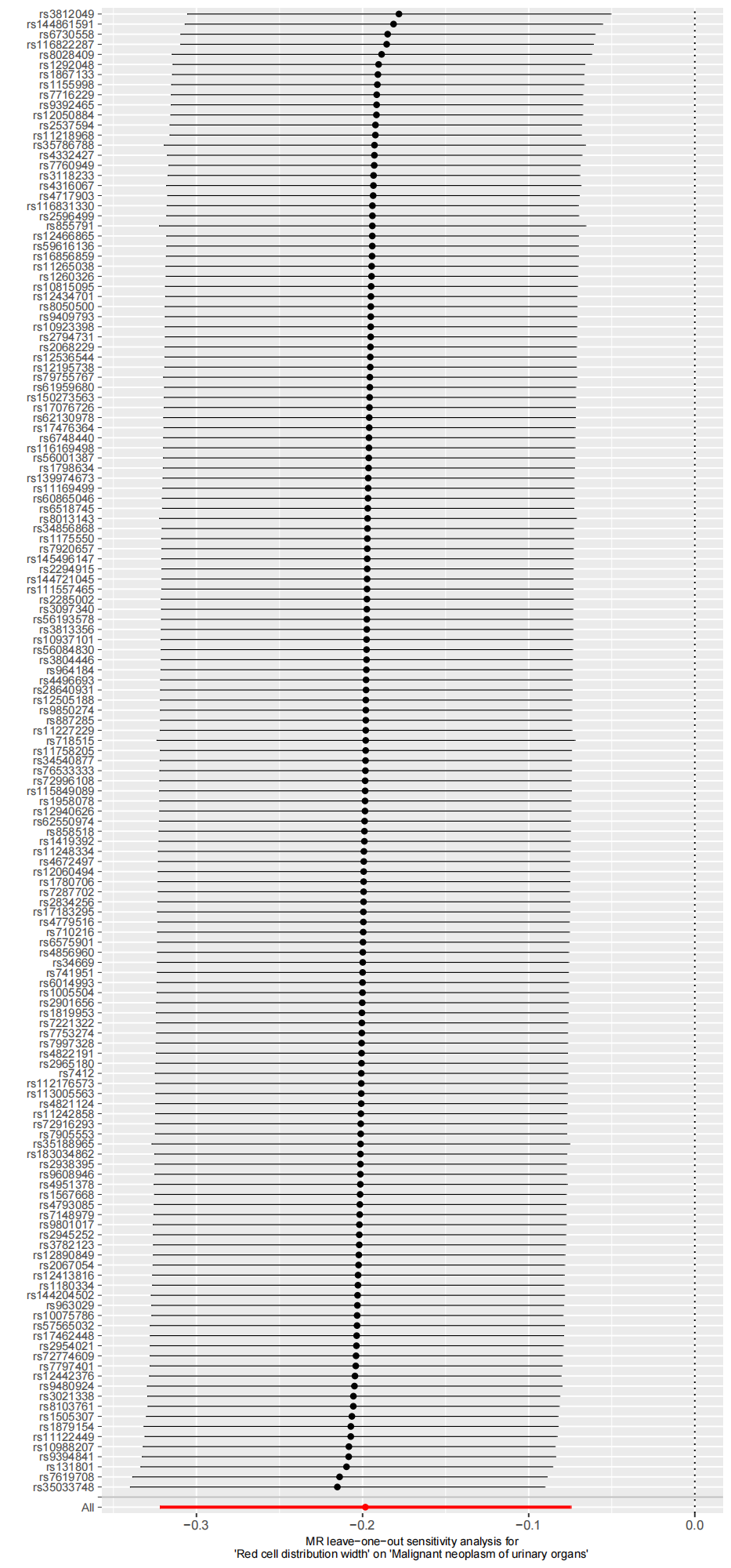

3) Scatter plot

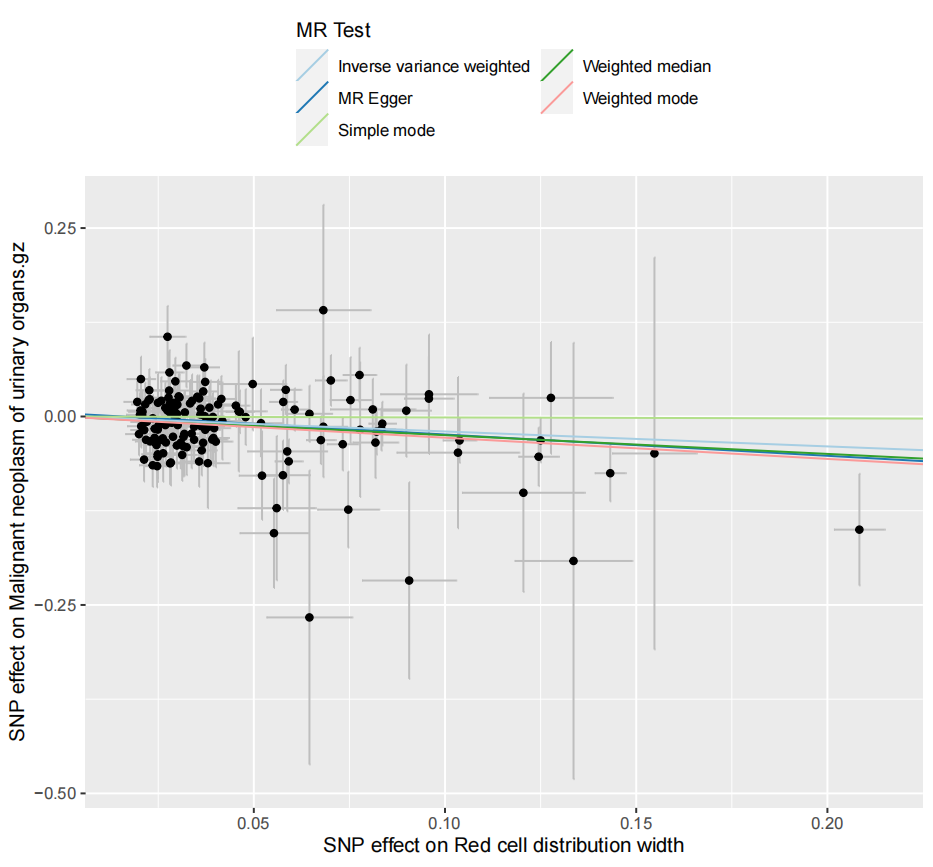

4) Funnel plot

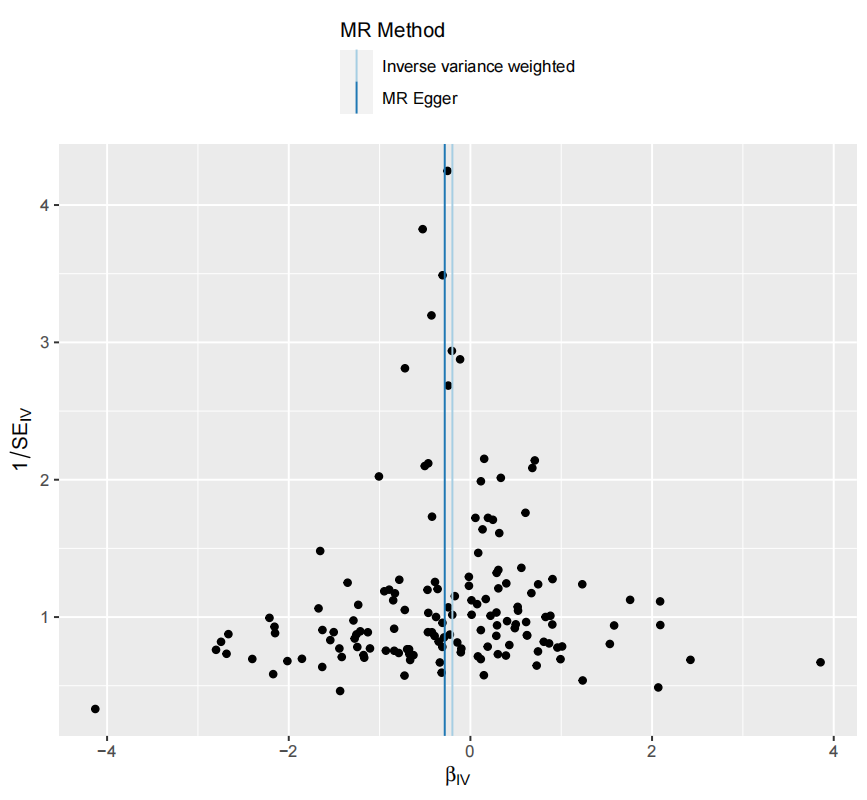

### Supplementary Figure 12. Genetic association of White blood cell count with Malignant neoplasm of oral cavity

1) Forest plot

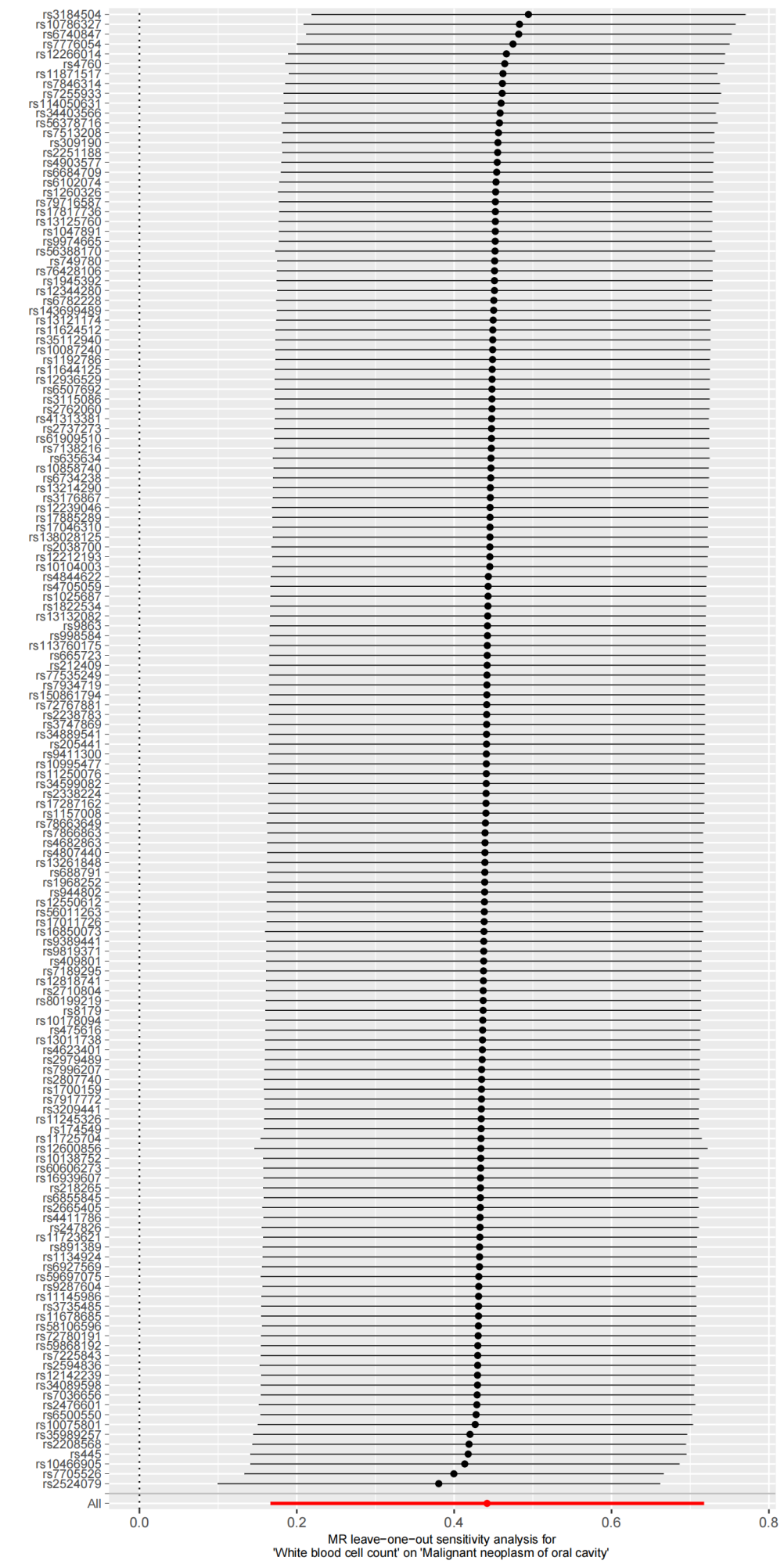

2) Leave-one-out plot

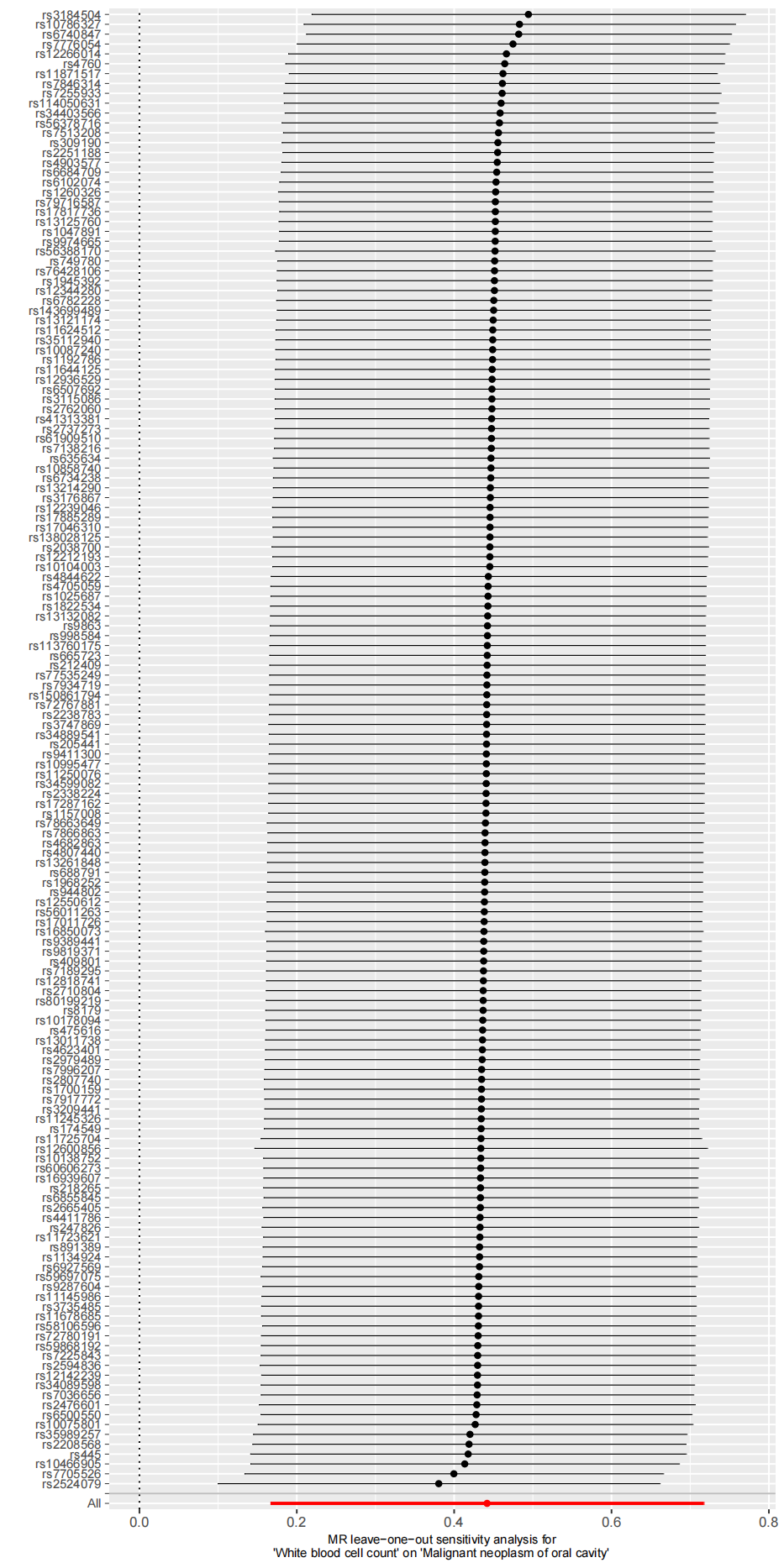

3) Scatter plot

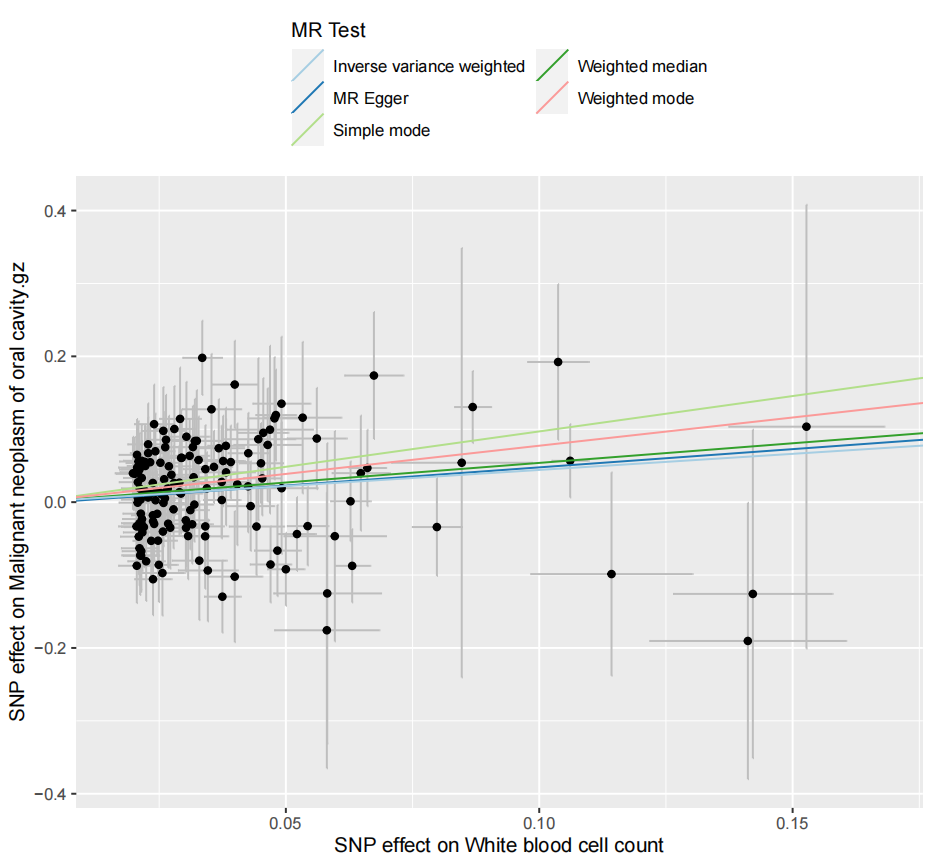

4) Funnel plot

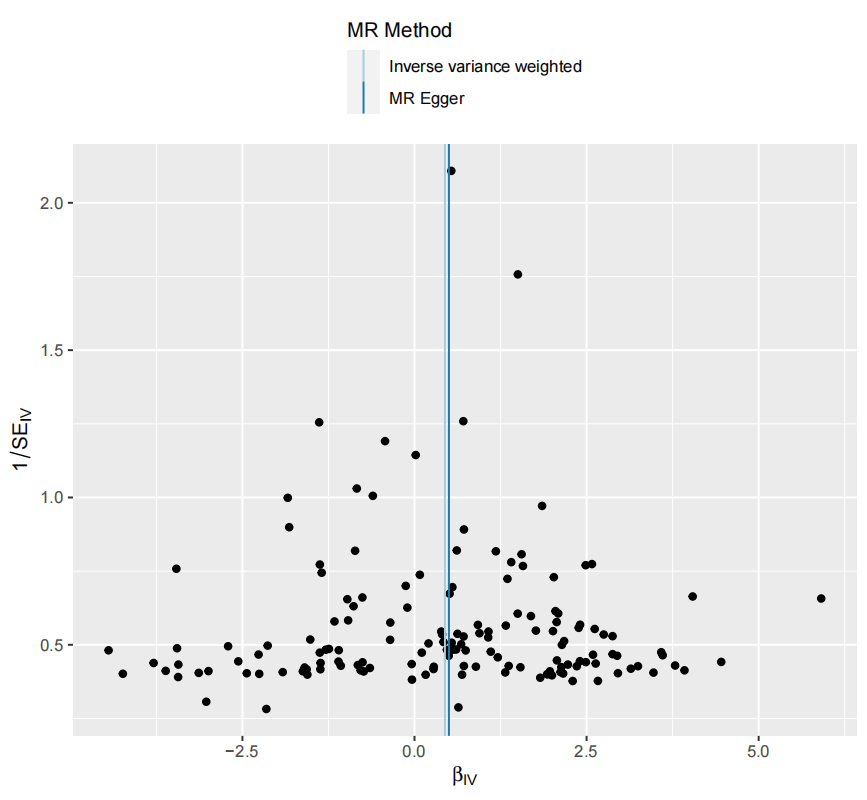

### Supplementary Figure 13. Genetic association of High light scatter reticulocyte count with Malignant neoplasm of stomach

1) Forest plot

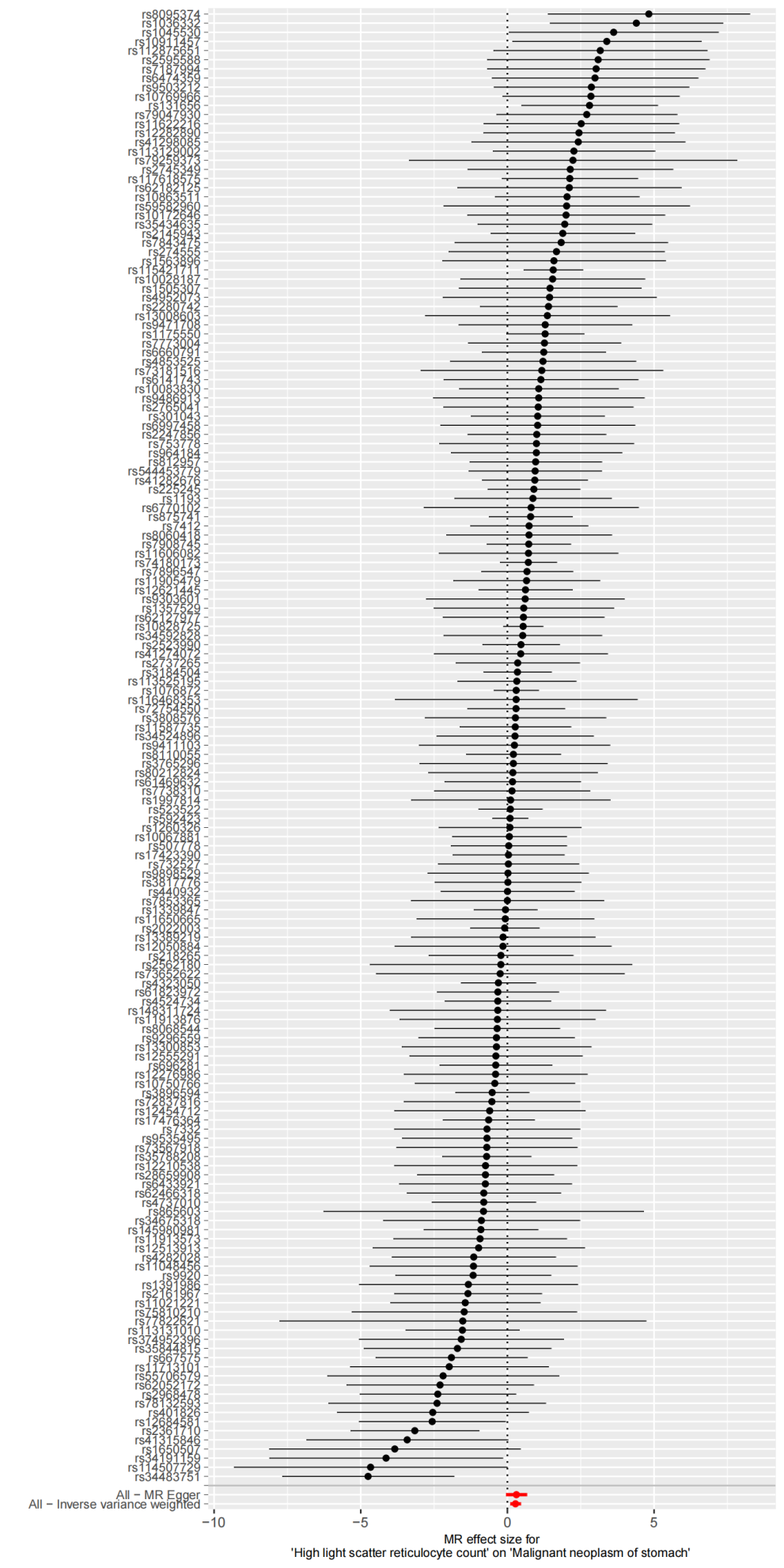

2) Leave-one-out plot

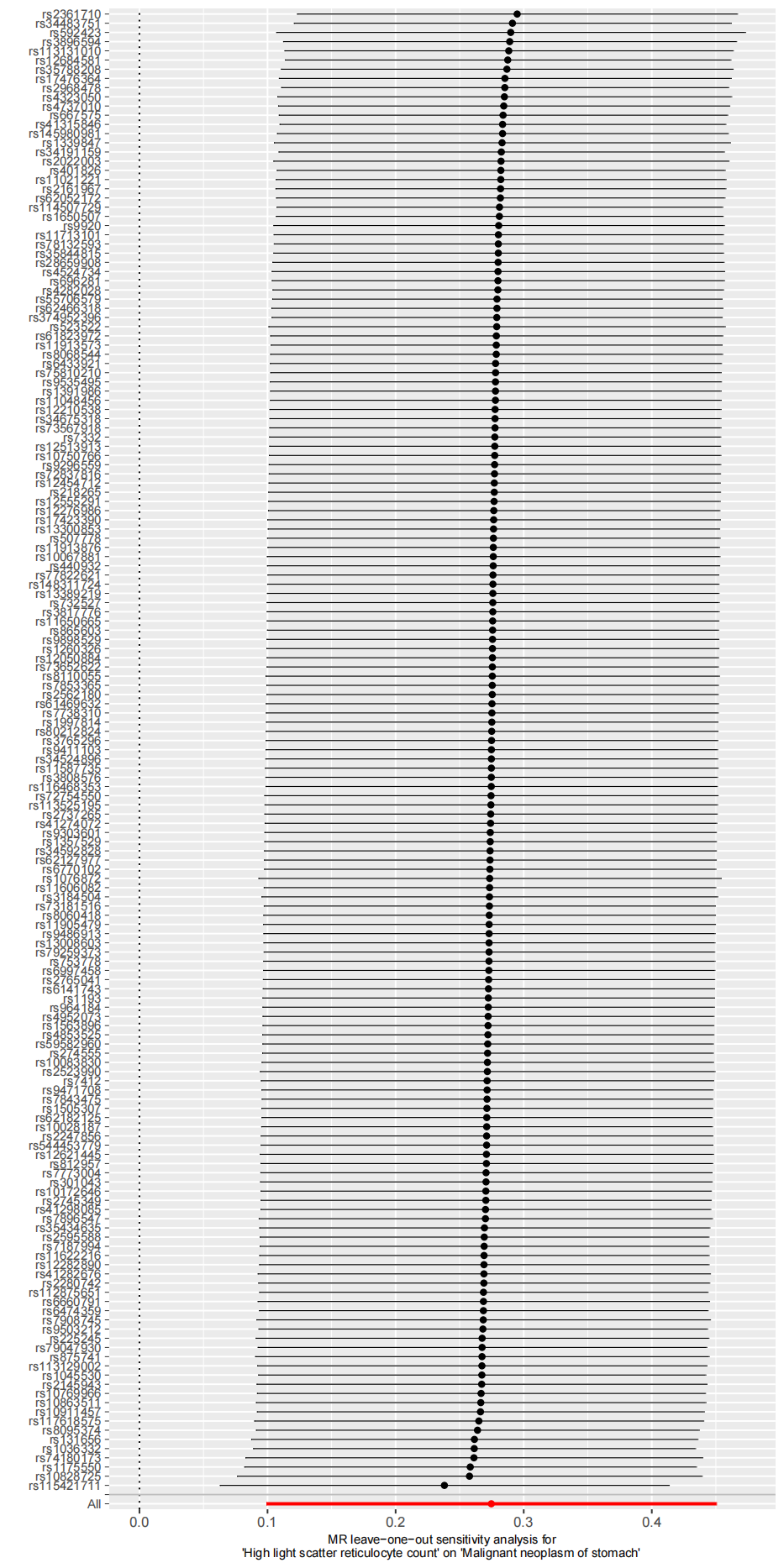

3) Scatter plot

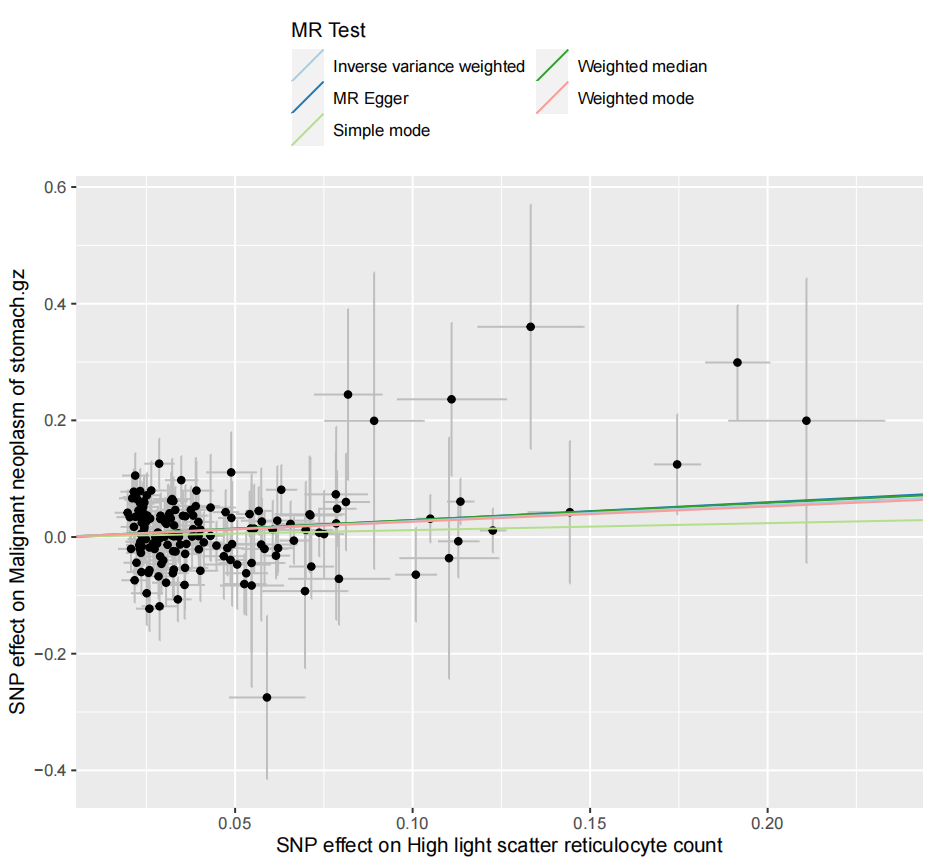

4) Funnel plot

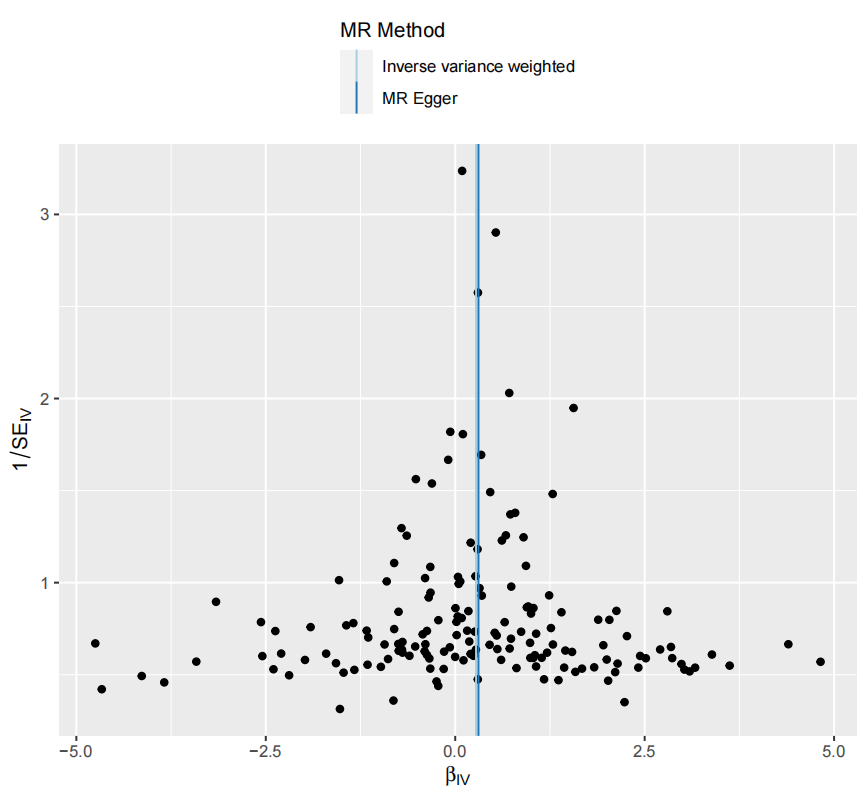
